# AccessPD - a ‘next generation’ registry to accelerate Parkinson’s disease research

**DOI:** 10.1101/2023.09.26.23295976

**Authors:** Yun-Hsuan Chang, Maria Teresa Periñan, Matt Wilson, Alastair J Noyce

## Abstract

**Objective:** To create a registry of patients with Parkinson’s disease (PD) and a rich database of PD-relevant information that can be used to stratify participants for precision opportunities.

**Background:** Recruitment to studies and trials is a major rate-limiting factor in PD research. Participants from lower socioeconomic backgrounds or geographically remote areas often have restricted access to clinical research opportunities. AccessPD is a novel platform that aims to accelerate progress of PD research.

**Design/Methods:** Potential participants are identified using electronic health records (EHRs) held by their primary care providers. They are contacted via a text message with an individualized link to the study portal. Electronic patient-reported outcomes (ePRO) are collected via regular online questionnaires and integrated with existing EHR.

**Results:** 200 participants were recruited within the first 6 months, with an average age of 70.8 years at the time of enrollment. When re-engaged, 191 participants answered the follow-up questionnaire. Here, to showcase the potential of AccessPD, we described the most common diagnoses before and after the diagnosis of PD, the most commonly prescribed drugs, and we used a case study to demonstrate how precision opportunities for research can be created by identifying participants who could benefit from device-aided therapies using consensus criteria.

**Conclusions:** AccessPD will enroll a minimum of 2000 patients. Early-stage analysis using ePROs and EHR data demonstrated AccessPD’s unique ability to link different data sources that could be used to stratify patients for longitudinal observational studies or recruit patients into clinical trials most suited to them.

## Introduction

Parkinson’s disease (PD) is a complex neurological condition with a broad range of clinical symptoms, which is influenced by both genetic and environmental factors ^1^. Management of PD calls for individualized pharmacological and non-pharmacological treatment. More research is needed to understand PD etiology, sub-types, progression rates and phenotype-genotype correlations in order to optimize management ^2,3^. Two needs for PD research are: 1) efficient ways to improve rate of participant recruitment, and 2) access to high-quality data from a diverse and representative sample of the population with PD. Inequity in access to research opportunities due to socioeconomic or geographical factors continues to bias our understanding of PD causation, manifestations, treatment adherence and response, and limits the generalisability of findings ^4^.

The widespread adoption of electronic health record (EHR) systems and increasing use of EHR data for secondary use (e.g. research) presented an opportunity to redesign a next-generation disease registry that is dynamic, interoperable and has the ability to effectively connect and combine data from different sources ^5^. This differs from traditional registries which require collaboration between large medical centers and active involvement of the clinicians to identify and collate cases ^6^.

Here, we describe AccessPD, a registry that aims to accelerate PD research by supporting participant enrollment and facilitating the collection of longitudinally-linked data for patient stratification. The system utilizes EHR data collected at the point of care at primary care practices across England to identify potential participants with a confirmed diagnosis of PD. Once a patient is contacted and consented into the registry, electronic patient-reported outcomes (ePRO), which are key indicators for disease progression and management, are collected via regular online questionnaires and integrated with existing EHR data, together with genetic or biomarker data that are obtained from home testing. Approved researchers can take advantage of the large, growing database and conduct novel studies to further our understanding of the disease. Partners wishing to validate devices or enroll participants to clinical trials can use AccessPD to recruit highly-stratified patients.

The remote nature of the registry ensures that participation is accessible to a more diverse population with PD than is typically seen in research studies.

This report summarises the recruitment of the first 200 patients to the AccessPD registry, and showcases the type of data that can be curated from re-engagement with questionnaires and from the EHR. We then show how this information can be used to stratify participants for precision research opportunities, using a specific case study to identify candidates with motor fluctuations to enroll in trials of device-aided therapies.

## Methods

### Engagement, Recruitment and high-level Design of AccessPD

AccessPD recruits patients via a digital platform powered by uMedeor LTD (uMed), a life sciences research organization that acts as a data processor for a network of primary care general practitioners (GPs) across England. uMed enables GPs to engage in research studies by automating the identification of potential study participants, a time-consuming step for the clinical staff, and facilitating participant engagement on behalf of the GPs. The registry is run and maintained by Cohort Science, a clinical research organization, in collaboration with researchers from Queen Mary University of London (QMUL). Figure 1 demonstrates the high-level design of the data ecosystem.

**Figure 1.**
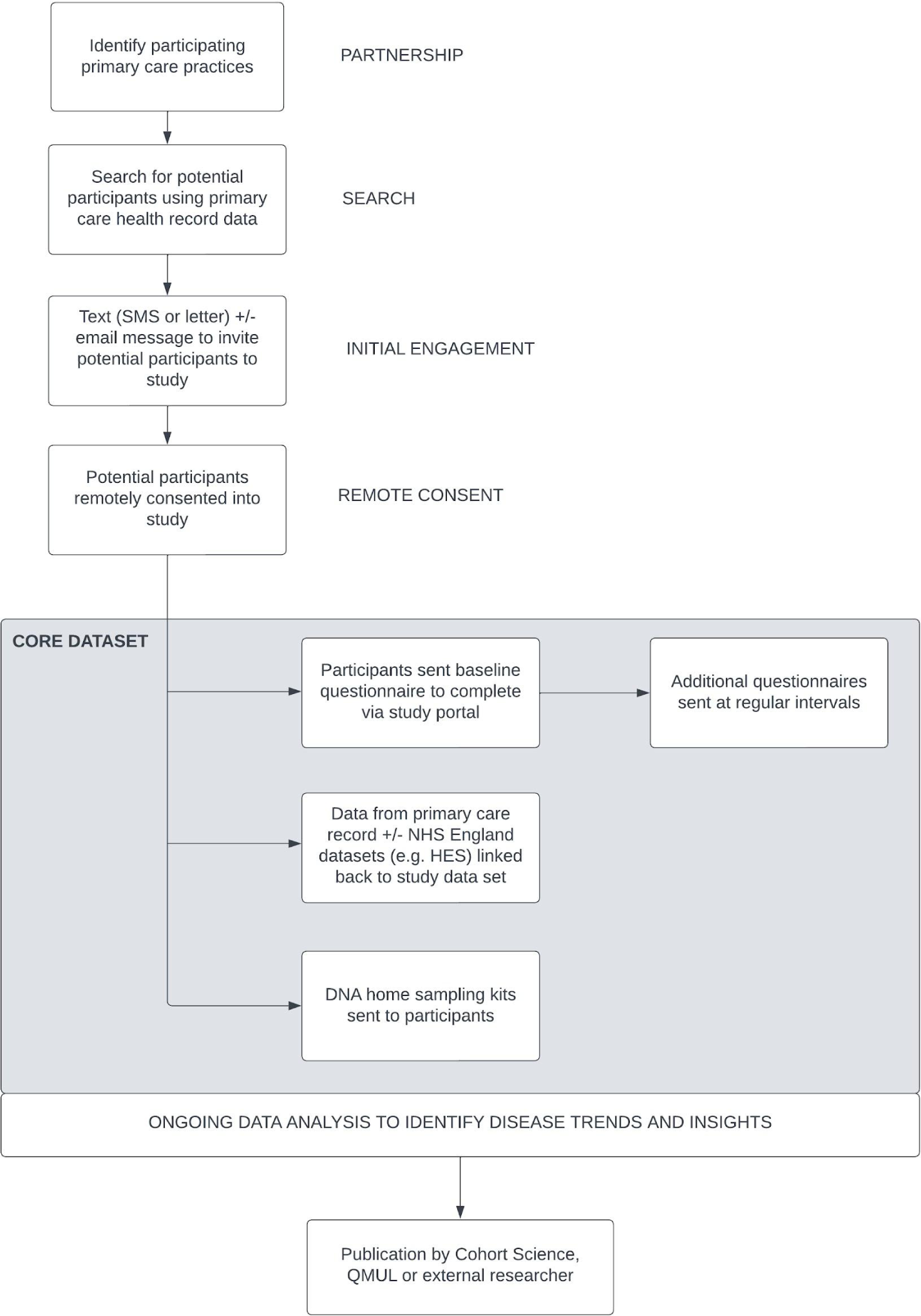
Design of AccessPD. Through partnerships with primary care providers, uMed acts on behalf of the GPs to identify potential candidates for AccessPD. Interested participants are engaged and consented remotely. NHS, National Health Services; HES, Hospital Episode Statistics; QMUL, Queen Mary University of London.

Patients with a coded diagnosis of PD – which is predominantly represented by the Systematised Nomenclature of Medicine Clinical Terms (SNOMED-CT) code 49049000 in primary care EHR – were identified as potential participants (additional diagnostic codes are listed in Supplementary Table 1). As PD is primarily managed in secondary or tertiary care in England, diagnosis records might be missing in primary care EHR systems. To mitigate the challenge of undercoding in primary care data, patients with a prescription of one or more anti-parkinsonian drugs were also identified as a subcohort of potential participants (Supplementary Table 2). Participants had to be above the age of 18, with capacity to give consent and have regular access to the internet or telecommunications.

With approval from participating GPs to engage these patients, uMed sends out a letter to inform them of the study opportunity, followed by a text message (either SMS or Email message) with an individualized link to the study portal. Once eligibility is established, participants give their consent to join the registry by going through a series of interactive questions. Alternatively, those less familiar with remote studies can request assistance from a study nurse who will go through the consent questions on the telephone with them and record answers in the database.

Incorporation of hospital episodes and electronic prescribing data from secondary and tertiary care is planned in the long-term design. In addition, the database will include genetic data and biomarker results from home testing kits. Approved researchers, affiliated or not with Cohort Science and QMUL, can access this resource via a secure web portal and conduct PD-relevant research. All participants will be informed of any use of their data in research and have the opportunity to opt out, should they wish to.

While maintaining data security and privacy, participants in the registry will be regularly informed of new study outcomes and opportunities via newsletters and re-consented for changes in study or privacy requirements, if necessary.

33 GP practices across England were involved in the project at launch in September 2022. The number of participating practices increased to 51 by the time the first 200 participants were recruited. All practices use either EMIS Health (90.2%) or SystemOne (9.8%), two of the main primary care EHR systems in the UK.

### Demographic data at baseline

Data such as age at enrollment, average years since PD diagnosis, ethnicity and gender were extracted from questionnaire-responses of the first 200 participants. Using their postcodes stored in EHR records, and the 2019 English deprivation data published by the Department for Levelling Up, Housing and Communities in the UK, we extracted the indices of multiple deprivation (IMD) for the 193 postcode areas where the participants reside. IMD is a score based on a number of socio-economic domains such as income, employment, education and health to represent relative deprivation of each small area in the UK ^7^. Descriptive data are presented as the mean and 95% confidence interval (CI) for parametric data. Categorical data are presented as proportions.

### Collecting ePROs and linking data

Consented participants received a baseline questionnaire immediately after enrollment that collected information on their demographics and current symptoms. Additional questionnaires were sent out at regular intervals. The first follow-up questionnaire consisted of 15 multiple-choice questions (see Supplementary Table 3) was sent out in March 2023 to the first 197 participants.

To create a linked database, primary care EHR data of the consented participants were incorporated into the registry to provide a longitudinal view of the medical history. Information was extracted separately from EMIS and SystemOne. EMIS Data Extraction Service was used for acquiring data from practices using the EMIS Health system whilst a proprietary tool developed by uMed was used to obtain data from collaborating SystemOne practices.

### Data linkage and processing

To demonstrate the ability of AccessPD in providing clinical insights, we conducted several analyses that utilized both ePROs and EHR data.

#### 1. Reliability of date of diagnosis

We compared the self-reported date of diagnosis with the diagnosis records of the first 200 participants extracted from primary care EHR. Inaccuracy in recall or delay in entering PD diagnosis into primary care EHR could lead to minor discrepancies between recalled and recorded date of diagnosis. As PD is a chronic disease and some participants only remember the closest month of diagnosis, we considered a difference of less than 12 months between the two dates as acceptable. Participants identified by their prescription records alone were excluded from this comparison because the first appearance of a PD medication prescription has limited relevance in determining the date of PD diagnosis if no treatment was commenced immediately.

#### 2. Symptoms/diagnoses prior and after PD diagnosis

By analyzing EHR observation records, we created a list of the most commonly used diagnostic codes among the participants before and after their diagnosis of PD. This analysis serves to provide a general insight into EHR data of the participants by looking at the codes that were most frequently applied during their encounters with their primary care providers.

To create a dataset for observation records, we first extracted the EMIS (carerecord_observation) and SystemOne (srcode) records separately and then joined the two tables using SNOMED-CT identifiers. The records were divided into two different datasets that represent all observation codes prior to and after the self-reported year of diagnosis for each individual patient.

Since the observation records contain more than just diagnoses and symptoms and certain diagnoses are represented by multiple codes (e.g. both “asthma” and “annual review of asthma” indicate the presence of an asthma diagnosis), the list of the most commonly used codes was reviewed by a clinician to keep only the relevant codes. Codes that represent administrative information or laboratory tests were discarded and not included in the ranking (see Supplementary Table 4 for codes discarded and Supplementary Table 5 for codes used in querying each symptom/diagnosis).

#### 3. Top 20 current medications at the time of enrollment

As with the observation records, we joined the prescription records of EMIS (Prescribing_DrugRecord) and SystemOne (SRPrimaryCareMedication) by dm+d codes that are common to both systems. Only records from 2021 on were taken into account. After removing duplicated drug prescriptions for each individual, we listed out the 20 most commonly prescribed drugs among the participants at the point of enrollment.

#### 4. Case study - identifying participants with motor fluctuations for device-aided therapy

Participants who had either been prescribed levodopa five times daily *or* had self-reported periods of ‘off’ time *or* dyskinesia were identified based on EHR records and answers in the questionnaires, respectively. This is in accordance with the ‘5-2-1’ screening criteria that is used to identify patients with advanced PD that may benefit from device-aided therapies ^8,9^. We then looked at EHR prescriptions to see if they had previously been treated with dopamine agonists, MAO-B inhibitors or COMT inhibitors. We used this case study to show how AccessPD could be used for a specific indication or to stratify patients for precision opportunities, such as identifying patients with complex PD who might be suitable for device-aided therapies.

## Results

### Initial engagement and baseline questionnaire

We identified a total of 1676 (0.27%) PD patients out of 628,610 unique patients registered with the first 51 participating GP practices, as of the 1st March 2023. Of the 1676 patients contacted for initial engagement, 404 (24.1%) responded by clicking on the link included in the text invitation and 232 (13.8%) participants completed all the consent questions. 200 consented and agreed to take part in AccessPD. The 200th patient with PD was recruited into AccessPD 182 days after the launch of the study.

Of the first 200 participants, 43.0% were female and 57.0% were male (Table 1). The most commonly reported symptoms were tremor (78.0%), muscle stiffness (64.5%), slowness of movement (58.5%), fatigue (60.0%), sleep disorder (49.0%) and gait problems (45.0%). A family history of PD was reported in 19.0%. Two (1%) participants had undergone deep brain stimulation.

**Table 1.** Participant demographic data and self-reported symptoms for the first 200 participants.

| Characteristics |  | Participants (n= 200) |
| --- | --- | --- |
| Average age (years) at diagnosis (95%CI) |  | 65.6 (64.2, 67.0) |
| Average years since diagnosis (95%CI) |  | 5.2 (4.5, 5.9) |
| Average age (years) at enrollment (95%CI) |  | 70.8 (69.5, 72.1) |
| Age at enrollment (years) | < 40 | 1 (0.5%) |
|  | 40-49 | 5 (2.5%) |
|  | 50- 59 | 13 (6.5%) |
|  | 60-69 | 56 (28.0%) |
|  | 70-79 | 97 (48.5%) |
|  | > 80 | 28 (14.0%) |
| Sex at birth, n(%) |  |  |
|  | Female | 86 (43.0%) |
|  | Male | 114 (57.0%) |
| Self-reported ethnicity, n (%) |  |  |
|  | White (including English, Welsh, Scottish, Northern Irish or Irish and any other White background) | 189 (94.5%) |
|  | Non-white (including Asian, black, mixed, other and prefer not to say) | 11 (5.5%) |
| Index of Multiple Deprivation of the postcode areas where the participants live |  |  |
|  | *Index of Multiple Deprivation Decile* | *Number of postcode areas, n(%)* |
|  | 1-2 | 24 (12.4%) |
|  | 3-4 | 35(18.1%) |
|  | 5-6 | 17 (8.8 %) |
|  | 7-8 | 44 (22.8%) |
|  | 9-10 | 73 (37.8%) |
| Most common current symptoms, n (%) |  |  |
|  | Tremor | 156 (78.0%) |
|  | Muscle stiffness | 129 (64.5%) |
|  | Fatigue | 120 (60.0%) |
|  | Slowness of movement | 117 (58.5%) |
|  | Problem sleeping | 98 (49.0%) |
|  | Problem walking | 90 (45.0%) |
| Family History of PD, n (%) |  |  |
|  | Yes | 38 (19.0%) |
|  | No | 162 (81.0%) |
| Impact of PD on movement, n (%) |  |  |
|  | No impact of motor symptoms or symptoms affect one side of the body | 100 (50.0%) |
|  | Symptoms affect both sides of the body | 78 (39.0%) |
|  | Assistance in daily activities needed | 22 (11.0%) |
| History of Deep Brain Stimulation, n (%) |  |  |
|  | Yes | 2 (1.0%) |
|  | No | 198 (99.0%) |

The average age at the time of recruitment was 70.8 years. The youngest participant was 39-year-old and the oldest was 90-year-old. 18 (9.0%) participants were diagnosed before the age of 50. With an average number of years since diagnosis of 5.2 years, the participant with the longest history of PD was diagnosed 31 years ago, and 27.5% of participants were diagnosed within the past 24 months.

50% of the participants reported they had either no PD motor symptoms or their symptoms were confined to only one side of the body. 39% of participants reported that both sides of their bodies were affected by PD or they struggled with walking and balance. 11% of participants need assistance with activities of daily living.

Analysis of the IMD scores showed that >30% of participants lived in the lowest two quintiles of deprivation in England (IMD 1-4). When asked about their ethnicity, the majority (94.5%) of the participants identified as white, with the remainder disclosing that they were Asian, mixed, black or preferred not to answer.

### First engagement questionnaire

A total of 191 registered participants answered the first follow-up questionnaire, 55 of them required assistance from a nurse via telecommunication to enter their responses. The survey results showed that 84.3% of the participants were comfortable with using mobile technology (laptop, tablet, computer or mobile phone) as the primary way for receiving communication. 78.5% of the participants have not participated in any form of PD-related research. Of those who had research experience in the past, the most common type was questionnaire-based research (52.2%), followed by clinical research that required trial site visits (21.3%) and research involving a medical device or testing kit (14.8%).

When asked about the symptoms that most negatively impacted quality of life, the most common answers were slowness of movement or loss of dexterity (51.3%), tremor (50.8%), walking problems such as abnormal gait, difficulty walking and falls (40.8%), fatigue (40.8%) and muscle stiffness (40.3%) (see Figure 2). Fatigue (22.0%), tremor (20.9%), walking problems (20.4%), bladder or bowel function disorders (20.4%) or slowness of movement (19.4%) were among the most common symptoms that were least controlled by their current medication. In general, 64.9% of participants said they were satisfied with their current therapy and management of PD.

**Figure 2.**
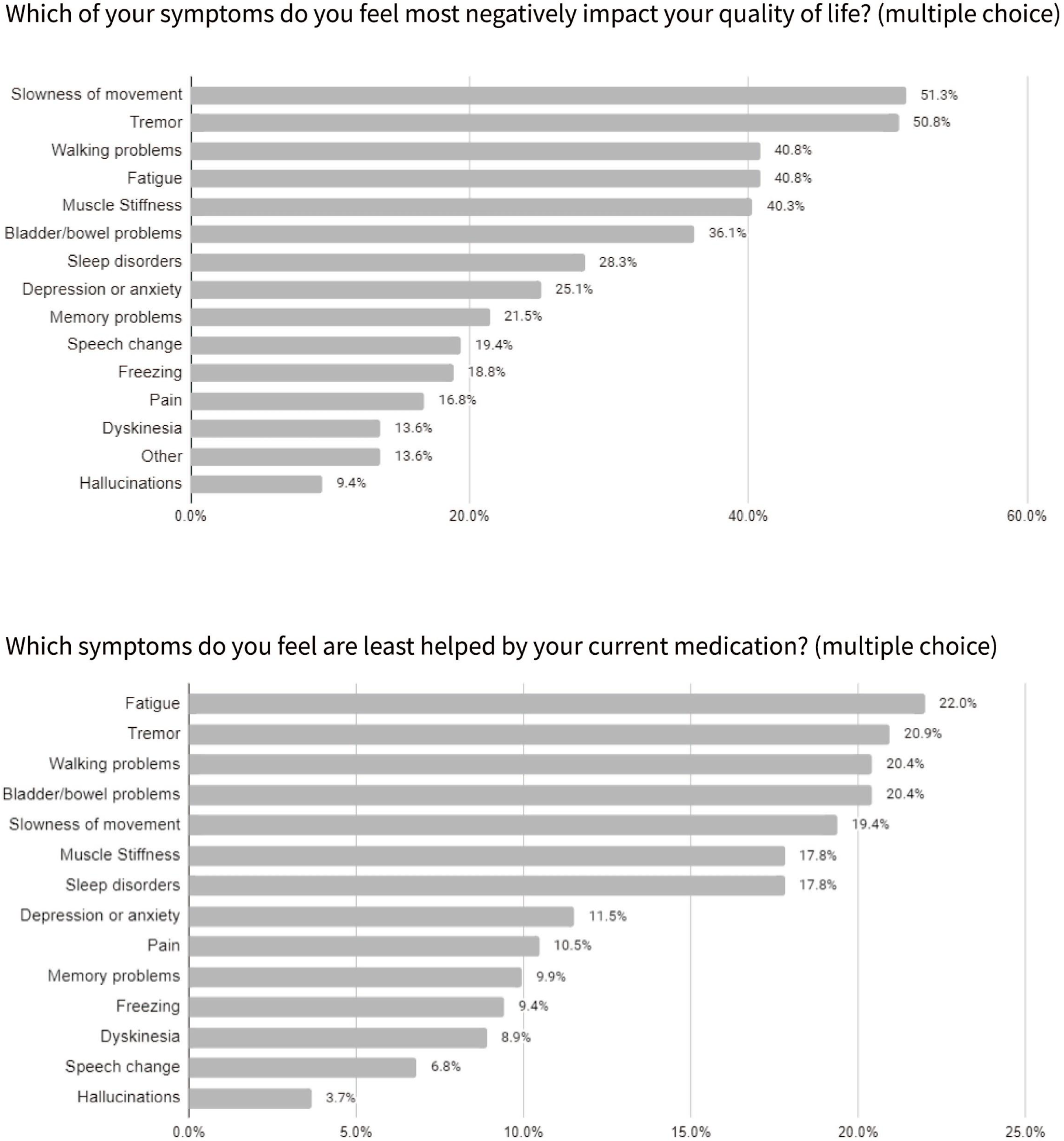
Bar charts showing symptoms that most negatively impact participants’ quality of life and symptoms that are least helped by their current medications.

36.8% of participants reported suffering from wearing ‘off’ periods. The remaining participants had either not noticed any ‘off’ periods or were unsure about this, and 6 (3%) responded that they were not yet on PD medication. We asked if participants were aware of device-aided therapy for PD. Only 6 out of 191 (3%) participants were aware of deep brain stimulation. One participant knew about apomorphine infusion or subcutaneous apomorphine, and one had heard about levodopa-carbidopa intestinal gel.

One of the 191 participants who responded to the follow-up questionnaire reported having multiple system atrophy-parkinsonian type (MSA-P) and was excluded from further analysis.

### Analysis of ePROs and EHR data

#### 1. Comparing date of diagnosis

83.9% of participants had a coded diagnosis of PD that first appeared within one year of the self-reported date of diagnosis (Supplementary Table 6). In most instances (80.2%) the self-reported date appeared earlier than the first EHR record of PD.

#### 2. Co-existing diagnoses prior to and after confirmation of PD

Table 2 summarises the most common diagnoses extracted from EHR records of the 190 participants, before and after their PD diagnosis. Musculoskeletal conditions, hypertensive disorder, depression and anxiety, urinary tract infection, skin conditions, haemorrhoids or constipation, and respiratory tract infection are among the most frequent reasons for a GP consultation both before and after the diagnosis. 14.2% of patients had a record indicating diabetes after PD diagnosis (versus 5.3% before the diagnosis).

**Table 2.** Most common co-existing diagnoses of the participants, before and after their PD diagnosis. The equal sign in the ranking column indicates the same code frequency.

| Before diagnosis of PD |  |  | After diagnosis of PD |  |  |
| --- | --- | --- | --- | --- | --- |
|  | Diagnosis/Symptom | No. of patients (%) |  | Diagnosis/Symptom | No. of patients (%) |
| 1 | Joint pain & Musculoskeletal disorders | 75 (39.5%) | 1 | Joint pain & Musculoskeletal disorders | 68 (35.8%) |
| 2 | Skin lesions, dermatitis & keratosis | 57 (30.0%) | 2 | Hypertensive Disorder | 61 (32.1%) |
| 3 | Hypertensive Disorder | 32 (16.8%) | 3 | Depression & Anxiety | 32 (16.8%) |
| 4 | Headache/ Migraine | 28 (14.7%) | 4 | Urinary tract infection | 31 (16.3%) |
| =4 | Depression & Anxiety | 28 (14.7%) | 5 | Skin lesions, dermatitis & keratosis | 30 (15.8%) |
| 6 | Haemorrhoids or constipation | 25 (13.2%) | 6 | Diabetes | 27 (14.2%) |
| =6 | Abdominal Hernia | 25 (13.2%) | 7 | Haemorrhoids or constipation | 20 (10.5%) |
| 8 | Respiratory tract infection | 24 (12.6%) | 8 | Asthma | 19 (10.0%) |
| =8 | Dyspepsia/Indigestion/reflux | 24 (12.6%) | 9 | Respiratory tract infection | 19 (10.0%) |
| 10 | Urinary tract infection | 21 (11.1%) | 10 | Rheumatoid arthritis | 12 (6.3%) |

#### 3. Top 20 current medications at the time of enrollment

A similar analysis was performed to list the most commonly prescribed drugs in the EHR starting from 2021, one year prior to enrollment. Other than the COVID-19 vaccines and anti-parkinsonain drugs (Co-carelodopa, Rasagiline, Co-beneldopa, Sinemet), antibiotics (Amoxicillin, Flucloxacillin, Doxycycline), laxatives, antidepressants (Sertraline), and Atorvastatin, were among the most frequently prescribed drugs (see Table 3).

**Table 3.** 20 most commonly prescribed drugs in the EHR, starting from a year before the start of the enrollment.

| **Name of medication** | **Number of patients (%)** |
| --- | --- |
| COVID-19 mRNA Vaccine Pfizer-BioNTech BNT162b2<br>30micrograms/0.3ml dose concentrate for suspension for injection<br>multidose vials (Pfizer Ltd) | 121 (63.7%) |
| Co-careldopa 12.5mg/50mg tablets | 76 (40.0%) |
| COVID-19 Vaccine AstraZeneca (ChAdOx1 S [recombinant])<br>5x10,000,000,000 viral particles/0.5ml dose solution for injection<br>multidose vials (AstraZeneca UK Ltd) | 58 (30.5%) |
| COVID-19 Vaccine Spikevax 0 (Zero)/O (Omicron) 0.1mg/ml<br>dispersion for injection multidose vials (Moderna, Inc) | 54 (28.4%) |
| Comirnaty Original/Omicron BA.1 COVID-19 mRNA Vaccine<br>15micrograms/15micrograms/0.3ml dose dispersion for injection<br>multidose vials (Pfizer Ltd) | 43 (22.6%) |
| Co-careldopa 25mg/100mg tablets | 30 (15.8%) |
| COVID-19 mRNA (nucleoside modified) Vaccine Moderna<br>0.1mg/0.5ml dose dispersion for injection multidose vials<br>(Moderna, Inc) | 30 (15.8%) |
| Amoxicillin 500mg capsules | 29 (15.3%) |
| Sertraline 50mg tablets | 21 (11.1%) |
| Omeprazole 20mg gastro-resistant capsules | 21 (11.1%) |
| Flucloxacillin 500mg capsules | 20 (10.5%) |
| Rasagiline 1mg tablets | 19 (10.0%) |
| Co-beneldopa 12.5mg/50mg capsules | 18 (9.5%) |
| Atorvastatin 20mg tablets | 18 (9.5%) |
| Laxido Orange oral powder sachets sugar free (Galen Ltd) | 18 (9.5%) |
| Doxycycline 100mg capsules | 18 (9.5%) |
| Paracetamol 500mg tablets | 18 (9.5%) |
| Macrogol compound oral powder sachets NPF sugar free | 17 (9.0%) |
| Sinemet 12.5mg/50mg tablets (Merck Sharp & Dohme Ltd) | 17 (8.9%) |
| Fluad Tetra vaccine suspension for injection 0.5ml pre-filled syringes (Seqirus UK Ltd) | 16 (8.4%) |

#### 4. Case study - identifying patients that might be suitable for device-aided therapies

Based on EHR medication records, 19 (10%) participants were prescribed levodopa 5 times daily, while 56 (29%) participants reported dyskinesia in the baseline questionnaire and 70 (37%) participants answered yes to wearing ‘off’ symptoms in the follow-up questionnaire. This meant a total of 95 (50%) participants were either on levodopa five times daily *or* reported wearing ‘off’ *or* dyskinesia. Of these, 58 (31%) participants had been prescribed adjunctive PD medications from one of the three group groups DA, MAOi or COMT. 43 (23%) were prescribed at least one drug from one of the three groups, 12 (6%) have been prescribed drugs from two of the groups, whilst three participants (2%) had been trialed on drugs from all three classes of adjunctive treatment.

## Discussion

Here we describe the creation and launch of the AccessPD registry; a next-generation platform to accelerate PD research. The principal goal of AccessPD is to accelerate progress by providing access to opportunities for patients, access to patients for researchers, and access to data for the research community. The ability to re-engage AccessPD participants rapidly and create highly stratified groups of patients using EHR information and self-reported data, makes this accelerated pathway for research tangible. Integration of DNA and biomarker collection over the next couple of years and further growth of the registry are planned.

The successful recruitment of the first 200 participants to the registry within 6 months of launch demonstrates the capability of EHR data in supporting targeted trial recruitment. On average, 7.7 participants were recruited into AccessPD per week, which is higher than the weekly average of 4.9 participants based on analysis of three similar decentralized studies reported in a review conducted by Myers et al. ^10^. Interestingly, 62.5% of the first 200 participants were above 70 years of age and ∼80% had never been involved in research. This reflects the higher prevalence of PD among older patients but also suggests that older age is not a barrier to participating in a digital disease registry such as AccessPD. Although the IMD scores calculated using postcodes of the participants only serve as a proxy for measuring the relative deprivation, it indicates that participants were recruited from a wide range of socioeconomic backgrounds ^11^.

One of the limitations of traditional registries is the lack of ability to keep pace with changing requirements and the cumbersome nature of re-engaging and re-consenting participants at scale for collection of additional data points or dissemination of the latest PD-related study information^12^. AccessPD’s strength in efficiently re-engaging participants in this regard is evidenced by the speed and rate of response to our first follow-up questionnaire where 95.5% of participants submitted their answers within 2 weeks. This differentiates AccessPD from other research databases that solely support population health research and lack the ability to re-engage participants.

We envisage a huge range of possibilities for AccessPD, including: 1) researchers being able to engage participants with remote collections of data and samples (e.g. questionnaires, biospecimens), 2) rapid testing and validation of devices and software designed for patients with PD, 3) rapid recruitment to investigator-led and/or commercial clinical trials. We conducted four separate analyses to demonstrate both the integration of data and its validity. We observed reassuring results with respect to EHR date of diagnosis and self-reported diagnosis (>90% concordant within 0-2 years) and expected results for concurrent diagnoses and prescriptions held in the EHR.

We then used a case study focused on identifying those who might be suitable for either new research studies or who might be considered for a change in clinical management. We used consensus criteria (5-2-1 criteria) to identify 58 potentially eligible AccessPD participants.^9^ Of note, only 3% of AccessPD participants were previously aware of device-aided therapies. The ability to stratify patients precisely according to the inclusion and exclusion criteria of a given research opportunity offers unparalleled efficiency in recruitment from the perspective of the research sponsors and participants.

The AccessPD model disrupts the traditional, inefficient and costly model in which those that develop devices or drugs, go one-by-one to specialists in secondary care to find patients for their trials. By shifting the focus to primary care, stratifying patients, and engaging directly with those patients, much of the bias and exclusivity of research to date is eroded. This is demonstrated by the inclusion of older participants in AccessPD and the spread of participants across different multiple deprivation indices, factors that often lead to exclusion from previous research opportunities due to age or financial burden^13^.

### Limitations

To maximize sensitivity of our search algorithm, we utilized both diagnosis and drug codes for the identification of potential PD patients. A participant is then asked to confirm their PD diagnosis during the registration. The problem with this approach is two-fold: patients who are approached based on their prescription data alone, but who do not have PD, may perceive the invitation to the registry as intrusive. Furthermore, the medication algorithm does not identify early-stage patients with PD who are not on medication, which could skew the distribution of participants towards the later-stage patients.

Using EHR data for secondary research comes with intrinsic limitations such as data inconsistency, incompleteness and inaccuracy, as they are “by-products” of routinely collected data. Physicians might use different codes to represent the same disease, or code a symptom rather than a diagnosis during consultation, thus making it challenging to determine the most frequent concurrent diagnoses. Nevertheless, EHR data is valuable in providing researchers a general idea of co-morbidities which could be used to form the basis of new hypotheses. By linking ePROs and EHR data, AccessPD provides a unique way to cross-reference and validate information.

As mentioned above, the current distribution of participants across strata of deprivation is highly encouraging. However, the proportion of non-white participants in the registry remains relatively low. This could be attributed to factors such as language barriers and limited diversity in the background population of primary care providers involved in the pilot project, which are concentrated in the southeast and northwest England. Improving diversity in AccessPD remains a priority.

### Conclusions and future directions

The decentralized design of AccessPD enables study opportunities to be offered to participants that are often overlooked and aims to erode biases in participant selection and improve generalisability of results. More work will be done to understand the low uptake of AccessPD among ethnic minority groups and develop enhancement strategies.

Our current effort is focused on increasing the number of participants in AccessPD, collecting DNA and information from wearable devices, and seeking partners who wish to recruit study participants to drug trials or device validation studies. We also welcome researchers to access the existing data to tackle important research questions.

## Data Availability

The datasets generated and/or analyzed during the current study are not publicly available due to privacy restrictions but are available from the corresponding author on reasonable request.

## Acknowledgment

We would like to express our gratitude to all patients who participate in AccessPD and continue to make valuable contributions. Additionally, we extend our thanks to all primary care providers in the uMed network who support AccessPD.

## Author’s Roles

M.W. and A.J.N conceived and designed the study. M.W. and Y-H.C. carried out the implementation. Y-H.C and M.T.P. performed the analysis of the results and Y-H.C wrote the manuscript with support from A.J.N., M.T.P. and M.W. All authors contributed to the editing of the final manuscript.

## Disclosures

### Financial Disclosures for the previous 12 months

The authors declare that there are no additional disclosures to report.

## Reference

1. Del Rey NLG, Quiroga-Varela A, Garbayo E, et al. Advances in Parkinson’s Disease: 200 Years Later. Front Neuroanat. 2018;12. Accessed November 3, 2022. https://www.frontiersin.org/articles/10.3389/fnana.2018.00113

2. Emamzadeh FN, Surguchov A. Parkinson’s Disease: Biomarkers, Treatment, and Risk Factors. Front Neurosci. 2018;12. Accessed March 16, 2023. https://www.frontiersin.org/articles/10.3389/fnins.2018.00612

3. Lawton M, Ben-Shlomo Y, May MT, et al. Developing and validating Parkinson’s disease subtypes and their motor and cognitive progression. J Neurol Neurosurg Psychiatry. 2018;89(12):1279–1287. doi:10.1136/jnnp-2018-318337

4. Simonet C, Bestwick J, Jitlal M, et al. Assessment of Risk Factors and Early Presentations of Parkinson Disease in Primary Care in a Diverse UK Population. JAMA Neurol. 2022;79(4):359–369. doi:10.1001/jamaneurol.2022.0003

5. Jensen PB, Jensen LJ, Brunak S. Mining electronic health records: towards better research applications and clinical care. Nat Rev Genet. 2012;13(6):395–405. doi:10.1038/nrg3208

6. Wu AD, Wilson AM. Parkinson’s disease population-wide registries in the United States: Current and future opportunities. Front Digit Health. 2023;5:1149154. doi:10.3389/fdgth.2023.1149154

7. Payne R, Abel G. UK indices of multiple deprivation - A way to make comparisons across constituent countries easier. Health Stat Q Off Natl Stat. 2012;53:22–37.

8. Aldred J, Anca-Herschkovitsch M, Antonini A, et al. Application of the “5-2-1” screening criteria in advanced Parkinson’s disease: interim analysis of DUOGLOBE. Neurodegener Dis Manag. 2020;10(5):309–323. doi:10.2217/nmt-2020-0021

9. Antonini A, Stoessl AJ, Kleinman LS, et al. Developing consensus among movement disorder specialists on clinical indicators for identification and management of advanced Parkinson’s disease: a multi-country Delphi-panel approach. Curr Med Res Opin. 2018;34(12):2063–2073. doi:10.1080/03007995.2018.1502165

10. Myers TL, Augustine EF, Baloga E, et al. Recruitment for Remote Decentralized Studies in Parkinson’s Disease. J Park Dis. 12(1):371–380. doi:10.3233/JPD-212935

11. Strong M, Maheswaran R, Pearson T. A comparison of methods for calculating general practice level socioeconomic deprivation. Int J Health Geogr. 2006;5(1):29. doi:10.1186/1476-072X-5-29

12. Bellgard M, Beroud C, Parkinson K, et al. Dispelling myths about rare disease registry system development. Source Code Biol Med. 2013;8(1):21. doi:10.1186/1751-0473-8-21

13. Vaswani PA, Tropea TF, Dahodwala N. Overcoming Barriers to Parkinson Disease Trial Participation: Increasing Diversity and Novel Designs for Recruitment and Retention. Neurother J Am Soc Exp Neurother. 2020;17(4):1724–1735. doi:10.1007/s13311-020-00960-0

