## Supplementary Material for "AccessPD - a ‘next generation’ registry to accelerate Parkinson’s disease research"

**Supplementary Table 1. Diagnostic codes for Parkinson's disease.**
The following SNOMED-CT codes indicate the presence of a PD diagnosis and were used in the search for PD patients. Some patients might have multiple of these codes in their EHR records. The number of patients identified using each code only gives a rough estimate of their frequency of use. They do not represent the actual number of patients engaged because the database searched contained only anonymised data which means patients who had deceased or were deregistered from GP practices in the uMed's network were not excluded. Furthermore, as of March 2023, only 51 practices within uMed's network participated in the recruitment.

| **SNOMED ID** | **Description** | **Number of patients with this code** |
|---|---|---|
| 49049000 | Parkinson's disease | 4951 |
| 907151000000108 | Seen by Parkinson's disease service | 957 |
| 425390006 | Dementia associated with Parkinson's Disease | 452 |
| 862081000000106 | Referral to Parkinson's service | 337 |
| 924261000000104 | Referral to community Parkinson's disease clinical nurse specialist | 270 |
| 879471000000102 | Referral to community Parkinson's service | 58 |
| 515841000000104 | History of Parkinson's disease | 44 |
| 718685006 | Orthostatic hypotension co-occurrent and due to Parkinson's disease | 9 |
| 101421000119107 | Dementia due to Parkinson's disease | 8 |
| 341551000000108 | Cerebral degeneration in Parkinson's disease | 2 |
| 715345007 | Young onset Parkinson disease | 1 |

**Supplementary Table 2. Drug codes for anti-parkinsonian drugs including levodopa, dopamine agonists, monoamine oxidase-B (MAO-B) inhibitors, Catechol O-Methyltransferase (COMT) inhibitors.**

| SNOMED Code | Description |
|---|---|
| 323107008 | Product containing precisely benserazide 12.5 milligram and levodopa 50 milligram/1 each conventional release oral capsule |
| 323108003 | Product containing precisely benserazide 25 milligram and levodopa 100 milligram/1 each conventional release oral capsule |
| 323109006 | Product containing precisely benserazide 50 milligram and levodopa 200 milligram/1 each conventional release oral capsule |
| 323110001 | Co-beneldopa 12.5mg/50mg dispersible tablet |
| 323112009 | Co-beneldopa 25mg/100mg dispersible tablet |
| 323119000 | Product containing precisely carbidopa 12.5 milligram and levodopa 50 milligram/1 each conventional release oral tablet |
| 323208008 | Product containing precisely pramipexole dihydrochloride 88 microgram/1 each conventional release oral tablet |
| 323209000 | Product containing precisely pramipexole dihydrochloride 180 microgram/1 each conventional release oral tablet |
| 323210005 | Product containing precisely pramipexole dihydrochloride 700 microgram/1 each conventional release oral tablet |
| 377268007 | Product containing precisely carbidopa 10 milligram and levodopa 100 milligram/1 each conventional release oral tablet |
| 377269004 | Product containing precisely carbidopa 25 milligram and levodopa 100 milligram/1 each conventional release oral tablet |
| 377270003 | Product containing precisely carbidopa 25 milligram and levodopa 250 milligram/1 each conventional release oral tablet |
| 323184004 | Product containing precisely ropinirole (as ropinirole hydrochloride) 1 milligram/1 each conventional release oral tablet |
| 13639711000001102 | Ropinirole 2mg modified-release tablets |
| 39109211000001100 | Co-careldopa 50mg/200mg modified-release tablets |
| 11178911000001104 | Caramet 25mg/100mg CR tablets (Teva UK Ltd) |
| 5299611000001105 | Stalevo 100mg/25mg/200mg tablets (Orion Pharma (UK) Ltd) |
| 15440811000001105 | Stalevo 75mg/18.75mg/200mg tablets (Orion Pharma (UK) Ltd) |
| 5302211000001106 | Stalevo 150mg/37.5mg/200mg tablets (Orion Pharma (UK) Ltd) |
| 15440311000001101 | Stalevo 125mg/31.25mg/200mg tablets (Orion Pharma (UK) Ltd) |
| 13644811000001105 | Stalevo 200mg/50mg/200mg tablets (Orion Pharma (UK) Ltd) |
| 5298311000001106 | Stalevo 50mg/12.5mg/200mg tablets (Orion Pharma (UK) Ltd) |
| 19701511000001106 | Stalevo 175mg/43.75mg/200mg tablets (Orion Pharma (UK) Ltd) |
| 39734911000001107 | Co-careldopa 25mg/250mg tablets |
| 39734611000001101 | Co-careldopa 10mg/100mg tablets |

| | |
|---|---|
| 39734811000001102 | Co-careldopa 25mg/100mg tablets |
| 39734711000001105 | Co-careldopa 12.5mg/50mg tablets |
| 14054611000001101 | Co-careldopa 25mg/100mg/5ml oral solution |
| 11815111000001106 | Co-careldopa 25mg/100mg/5ml oral suspension |
| 8427211000001103 | Co-careldopa 12.5mg/50mg/5ml oral suspension |
| 8427611000001101 | Co-careldopa 6.25mg/25mg/5ml oral suspension |
| 39109011000001105 | Co-careldopa 25mg/100mg modified-release tablets |
| 391092110000011006 | Co-careldopa 50mg/200mg modified-release tablets |
| 10161711000001105 | Co-careldopa 5mg/20mg/1ml intestinal gel 100ml cassette |
| 27911000001105 | Madopar CR 125 capsules (Roche Products Ltd) |
| 379211000001105 | Madopar 250 capsules (Roche Products Ltd) |
| 129911000001109 | Madopar 125 capsules (Roche Products Ltd) |
| 73911000001102 | Madopar 62.5 capsules (Roche Products Ltd) |
| 570711000001108 | Madopar 125 dispersible tablets (Roche Products Ltd) |
| 192211000001104 | Madopar 62.5 dispersible tablets (Roche Products Ltd) |
| 562411000001105 | Sinemet 25mg/250mg tablets (Organon Pharma (UK) Ltd) |
| 790711000001101 | Sinemet 10mg/100mg tablets (Organon Pharma (UK) Ltd) |
| 236411000001107 | Sinemet 12.5mg/50mg tablets (Organon Pharma (UK) Ltd) |
| 826111000001108 | Sinemet CR 50mg/200mg tablets (Organon Pharma (UK) Ltd) |
| 210411000001104 | Sinemet Plus 25mg/100mg tablets (Organon Pharma (UK) Ltd) |
| 242011000001103 | Half Sinemet CR 25mg/100mg tablets (Organon Pharma (UK) Ltd) |
| 39707211000001101 | Co-beneldopa 25mg/100mg dispersible tablets sugar free |
| 39706911000001107 | Co-beneldopa 12.5mg/50mg dispersible tablets sugar free |
| 10161911000001107 | Duodopa intestinal gel cassette 100ml (Solvay Healthcare Ltd) \| |
| 3661711000001106 | Mirapexin 700microgram tablets (Boehringer Ingelheim Ltd) |
| 13127011000001109 | Mirapexin 350microgram tablets (Boehringer Ingelheim Ltd) |
| 3663011000001105 | Mirapexin 180microgram tablets (Boehringer Ingelheim Ltd) |
| 3663411000001101 | Mirapexin 88microgram tablets (Boehringer Ingelheim Ltd) |
| 16162611000001100 | Mirapexin 2.1mg modified-release tablets (Boehringer Ingelheim Ltd) |
| 16162911000001106 | Mirapexin 3.15mg modified-release tablets (Boehringer Ingelheim Ltd) |
| 18042311000001102 | Mirapexin 1.57mg modified-release tablets (Boehringer Ingelheim Ltd) |
| 18042611000001107 | Mirapexin 2.62mg modified-release tablets (Boehringer Ingelheim Ltd) |
| 16162011000001107 | Mirapexin 0.52mg modified-release tablets (Boehringer Ingelheim Ltd) |
| 323186002 | Product containing precisely ropinirole (as ropinirole hydrochloride) 5 milligram/1 each conventional release oral tablet |
| 323185003 | Product containing precisely ropinirole (as ropinirole hydrochloride) 2 milligram/1 each conventional release oral tablet |
| 323183005 | Product containing precisely ropinirole (as ropinirole hydrochloride) 250 |

| | |
|---|---|
| | microgram/1 each conventional release oral tablet |
| 13639911000001100 | Ropinirole 8mg modified-release tablets |
| 13639811000001105 | Ropinirole 4mg modified-release tablets |
| 13639711000001102 | Ropinirole 2mg modified-release tablets |
| 29184311000001107 | Ropinirole 3mg modified-release tablets |
| 29184411000001100 | Ropinirole 6mg modified-release tablets |
| 379611000001107 | ReQuip 2mg tablets (GlaxoSmithKline) |
| 481511000001109 | ReQuip 250microgram tablets (GlaxoSmithKline) |
| 923211000001100 | ReQuip 5mg tablets (GlaxoSmithKline) |
| 268111000001107 | ReQuip 1mg tablets (GlaxoSmithKline) |
| 3891211000001106 | ReQuip tablets starter pack (GlaxoSmithKline) |
| 3889511000001109 | ReQuip tablets follow on pack (GlaxoSmithKline) |
| 13628411000001102 | ReQuip XL 4mg tablets (GlaxoSmithKline) |
| 13628111000001107 | ReQuip XL 2mg tablets (GlaxoSmithKline) |
| 13628711000001108 | ReQuip XL 8mg tablets (GlaxoSmithKline) |
| 35935911000001108 | Rotigotine 4mg/24hours transdermal patches |
| 10142111000001106 | Rotigotine 8mg/24hours patches |
| 35936011000001100 | Rotigotine 6mg/24hours transdermal patches |
| 16683311000001101 | Rotigotine 1mg/24hours transdermal patches |
| 16665711000001104 | Rotigotine 3mg/24hours transdermal patches |
| 35935811000001103 | Rotigotine 2mg/24hours transdermal patches |
| 10129911000001102 | Neupro transdermal patches starter pack (Schwarz Pharma Ltd) |
| 10127311000001108 | Neupro 2mg/24hours transdermal patches (Schwarz Pharma Ltd) |
| 10128911000001105 | Neupro 6mg/24hours transdermal patches (Schwarz Pharma Ltd) |
| 10128311000001109 | Neupro 4mg/24hours transdermal patches (Schwarz Pharma Ltd) |
| 10129411000001105 | Neupro 8mg/24hours transdermal patches (Schwarz Pharma Ltd) |
| 16669211000001109 | Neupro 1mg/24hours transdermal patches (UCB Pharma Ltd) |
| 16664911000001101 | Neupro 3mg/24hours transdermal patches (UCB Pharma Ltd) |
| 5322411000001101 | Levodopa 150mg / Carbidopa 37.5mg / Entacapone 200mg tablets |
| 15457511000001109 | Levodopa 75mg / Carbidopa 18.75mg / Entacapone 200mg tablets |
| 5322511000001102 | Levodopa 50mg / Carbidopa 12.5mg / Entacapone 200mg tablets |
| 13648111000001109 | Levodopa 200mg / Carbidopa 50mg / Entacapone 200mg tablets |
| 5322311000001108 | Levodopa 100mg / Carbidopa 25mg / Entacapone 200mg tablets |
| 15457411000001105 | Levodopa 125mg / Carbidopa 31.25mg / Entacapone 200mg tablets |
| 19744111000001107 | Levodopa 175mg / Carbidopa 43.75mg / Entacapone 200mg tablets |
| 418125001 | Rasagiline 1mg tablets |
| 9362611000001103 | Azilect 1mg tablets (Teva UK Ltd) |

| | |
|---|---|
| 323158005 | Selegiline 5mg tablets |
| 323157000 | Selegiline 10mg tablets |
| 35938811000001107 | Selegiline 10mg/5ml oral solution |
| 323155008 | Selegiline 1.25mg oral lyophilisates sugar free |
| 32696511000001102 | Safinamide 50mg tablets |
| 32696411000001101 | Safinamide 100mg tablets |
| 323202009 | Entacapone 200mg tablets |
| 8487111000001103 | Entacapone 200mg/5ml oral suspension |
| 5322411000001101 | Levodopa 150mg / Carbidopa 37.5mg / Entacapone 200mg tablets |
| 15457511000001109 | Levodopa 75mg / Carbidopa 18.75mg / Entacapone 200mg tablets |
| 5322511000001102 | Levodopa 50mg / Carbidopa 12.5mg / Entacapone 200mg tablets |
| 13648111000001109 | Levodopa 200mg / Carbidopa 50mg / Entacapone 200mg tablets |
| 5322311000001108 | Levodopa 100mg / Carbidopa 25mg / Entacapone 200mg tablets |
| 15457411000001105 | Levodopa 125mg / Carbidopa 31.25mg / Entacapone 200mg tablets |
| 19744111000001107 | Levodopa 175mg / Carbidopa 43.75mg / Entacapone 200mg tablets |
| 375993004 | Tolcapone 100mg tablets |
| 33495211000001103 | Opicapone 50mg capsules |
| 33494911000001108 | Ongentys 50mg capsules (BIAL Pharma UK Ltd) |
| 323133000 | Amantadine 100mg capsules |
| 35899611000001103 | Amantadine 50mg/5ml oral solution sugar free |
| 35902411000001102 | Apomorphine 30mg/3ml solution for injection pre-filled disposable devices |
| 35902611000001104 | Apomorphine 50mg/5ml solution for injection ampoules |
| 35902311000001109 | Apomorphine 20mg/2ml solution for injection ampoules |
| 134614006 | Apomorphine 2mg sublingual tablets sugar free |
| 134615007 | Apomorphine 3mg sublingual tablets sugar free |
| 35902511000001103 | Apomorphine 50mg/10ml solution for infusion pre-filled syringes |
| 36441811000001103 | Apomorphine 30mg/3ml solution for injection cartridges |
| 36441211000001104 | Apomorphine 100mg/20ml solution for infusion vials |

**Supplementary Table 3. Template for the first follow-up questionnaire**

| Question |
|---|
| Q1: Do you prefer to answer the questionnaires that we send to you on your mobile, tablet, laptop or speaking over the phone ? |
| Mobile |
| Tablet |
| Laptop |
| Over the phone |
| Letter |
| Other (please specify) |
| Q2: Have you been offered the chance to participate in Parkinson's research before or is Access-PD the first opportunity? |
| ACCESS-PD is the first |
| 1-5 studies |
| >5 studies |
| other |
| Q3: If you have participated in Parkinson's research before, what type of research was this (you can select multiple options)? |
| Questionnaire-based |
| Involved a medical device or testing kit |
| Clinical trial completed at home only |
| Clinical trial completed involving travel (to a hospital or GP surgery) |
| I've never participated in Parkinson's research before AccessPD |
| Q4: Are you satisfied with your current medication / therapy for Parkinson's? |
| Yes |
| No |
| Other (please specify) |
| Q5: What would be your preferred method of taking your Parkinson's medication? |
| Pill once a day |
| Pill multiple times per day |
| Multiple pills multiple times per day |
| Skin patches |
| Injections |
| Infusions |
| Other (Please specify) |
| Q6: Which of your symptoms do you feel most negatively impacts your quality of life (you can select multiple options)? |
| Tremor |

| |
|---|
| Muscle Stiffness |
| Slowness of movement or loss of dexterity |
| Dyskinesia (uncontrolled, involuntary movement) |
| Walking problems (abnormal gait, difficulty walking, falls) |
| Speech change |
| Sleep disorders |
| Fatigue |
| Depression or anxiety |
| Memory problems |
| Hallucinations |
| Freezing (suddenly not be able to move forward for several seconds or minutes, feeling like your lower body is stuck) |
| Pain |
| Bladder or bowel problems |
| Other (please specify) |
| Q7: Which of your symptoms do you feel most negatively impacts your ability to carry out daily activities (you can select multiple options)? |
| Tremor |
| Muscle Stiffness |
| Slowness of movement or loss of dexterity |
| Dyskinesia (uncontrolled, involuntary movement) |
| Walking problems (abnormal gait, difficulty walking, falls) |
| Speech change |
| Sleep disorders |
| Fatigue |
| Depression or anxiety |
| Memory problems |
| Hallucinations |
| Freezing (suddenly not be able to move forward for several seconds or minutes, feeling like your lower body is stuck) |
| Pain |
| Bladder or bowel problems |
| Other |
| Q8: Which symptoms do you feel are best treated by your current medication (you can select multiple options)? |
| Tremor |
| Muscle Stiffness |
| Slowness of movement or loss of dexterity |

| |
|---|
| Dyskinesia (uncontrolled, involuntary movement) |
| Walking problems (abnormal gait, difficulty walking, falls) |
| Speech change |
| Sleep disorders |
| Fatigue |
| Depression or anxiety |
| Memory problems |
| Hallucinations |
| Freezing (suddenly not be able to move forward for several seconds or minutes, feeling like your lower body is stuck) |
| Pain |
| Bladder or bowel problems |
| Other (please specify) |
| Q9: Which symptoms do you feel are least helped by your current medication? |
| Tremor |
| Muscle Stiffness |
| Slowness of movement or loss of dexterity |
| Dyskinesia (uncontrolled, involuntary movement) |
| Walking problems (abnormal gait, difficulty walking, falls) |
| Speech change |
| Sleep disorders |
| Fatigue |
| Depression or anxiety |
| Memory problems |
| Hallucinations |
| Freezing (suddenly not be able to move forward for several seconds or minutes, feeling like your lower body is stuck) |
| Pain |
| Bladder or bowel problems |
| Other (please specify) |
| Q10: Do you suffer from on/off periods (medications wear off gradually or abruptly, resulting in the return of Parkinson’s symptoms)? |
| Yes |
| No |
| Other |
| Q11: If you answered ‘Yes’, does your medication do a good job of managing the number of on/off periods? |

| |
|---|
| Yes |
| No |
| I do not suffer from on/off periods |
| Other (please specify) |
| Q12: Do you do anything or take anything in addition to your medication to manage on/off periods? |
| Yes (please specify) |
| No |
| I do not suffer from On/Off periods |
| Other (please specify) |
| Q13: Have you switched medication that you take for Parkinson's in the past and if so, what reasons did you have for switching? |
| Medication did not improve symptoms (please specify the main symptom that medication did not help) |
| Side effects |
| Cost |
| Injection site reactions (if injected medication) |
| Other (please specify) |
| Q14: Have you ever been made aware of or been offered a device aided therapy by your physician? |
| Yes - deep brain stimulation |
| Yes - apomorphine infusion / sub-cutaneous apomorphine |
| Yes - levodopa-carbidopa intestinal gel |
| No |
| Not sure |
| Other |
| Q15: Do you have any agitators or specific tasks that cause your symptoms to increase? An example could be that flossing your teeth causes your tremor to increase. |
| Yes - Please specify |
| No |

**Supplementary Table 4. Codes discarded when analysing the most common co-exisiting diagnoses before and after the confirmation of Parkinson's disease.** These codes are discarded as they do not represent or indicate the presence of a medical diagnosis.

| **Before the confirmation of PD diagnosis** | |
|---|---|
| **SNOMED codes** | **Description** |
| 184229000 | Notes summary on computer |
| 965831000000103 | Implied consent for core Summary Care Record dataset upload |
| 773011000000101 | Patient allocated named accountable general practitioner |
| 313334002 | Blood sample taken |
| 908481000000105 | Informing patient of named accountable general practitioner |
| 266919005 | Never smoked tobacco |
| 182836005 | Telephone encounter |
| 279991000000102 | Administrative procedure |
| 185317003 | Medication review |
| 14734007 | Medication review done |
| 314530002 | Medication review with patient |
| 88551000000109 | Patient review |
| 763288003 | Result |
| 394617004 | British or mixed British - ethnic category 2001 census |
| 183049006 | Ex-smoker |
| 92391000000108 | Did not attend - no reason |
| 11816003 | Awaiting clinical code migration to EMIS Web |
| 8517006 | Preferred method of contact: unknown |
| 270426007 | SMS (short message service) text message sent to patient |
| 15728710000061005 | O/E - pulse rhythm regular |
| 185210004 | Failed encounter - message left on answer machine |
| 95324001 | SMS text message sent to patient |
| 185324002 | Patient mobile telephone number |
| 183093006 | Current non-smoker |
| 279039007 | Had a chat to patient |
| 185340004 | Discharge summary |
| 428481002 | Medication requested |
| 160618006 | Main spoken language English |
| 373942005 | Informed consent for procedure |
| 171071003 | Choice and booking enhanced services administration |

| | |
|---|---|
| 315570003 | Bowel cancer screening programme |
| 182771004 | FBC - full blood count |
| 222711000000102 | Exercise grading |
| 29857009 | NHS Health Check completed |
| 286901000000107 | Failed encounter |
| 523221000000100 | Letter from specialist |
| 30989003 | Letter encounter |
| 49727002 | Seen in hospital casualty |
| 45326000 | Telephone triage encounter |
| 59621000 | Advice on foreign travel |
| 160628002 | Administration |
| 308720009 | Letter from consultant |
| 225323000 | Patient's condition improved |
| 401267002 | Dietary advice |
| 416118004 | GFR (glomerular filtration rate) calculated by abbreviated Modification of Diet in Renal Disease Study Group calculation |
| 275693003 | Marital state unknown |
| 398838000 | Seen in GP's surgery |
| 1020291000000100 | Seen in dermatology clinic |
| 160504008 | Enjoys moderate exercise |
| 185202000 | Seen in orthopaedic clinic |
| 270422009 | Lifestyle education |
| 313204009 | Patient-initiated diet |
| 172630005 | Patient given advice |
| 160632008 | Serum lipids level |
| 160660005 | Mammography normal |
| 304507003 | Cervical smear - negative |
| 185252009 | Thyroid function test |
| 46825001 | Health education - exercise |
| 767357000 | White British |
| 168749009 | Bowel cancer screening programme faecal occult blood test normal |
| 185228002 | Enjoys light exercise |
| 1005661000000100 | Patient informed - test result |
| 167261002 | Urine glucose test negative |
| 165355002 | Systolic arterial pressure |
| 269958004 | Referral for further care |
| 315236000 | Depression screening using questions |
| 160631001 | Medication review of medical notes |

| | |
|---|---|
| 20135006 | Diastolic arterial pressure |
| 10168510000001005 | Sampling of cervix for Papanicolaou smear done |
| 183444007 | General practice physical activity questionnaire physical activity index: active |
| 60621009 | Body mass index |
| 93311000000106 | Smoking cessation advice |
| 200971000000100 | Seen in urology clinic |
| 366241000000106 | Education about alcohol consumption |
| 81680005 | Date records held from |
| 72313002 | General practice physical activity questionnaire physical activity index: moderately active |
| 161891005 | Letter encounter from patient |
| 162415008 | White British - ethnic category 2001 census |
| 162031009 | Liver function test |
| 165333005 | Urine protein test negative |
| 50417007 | Seen in cardiac clinic |
| 281078001 | MED3 - doctor's statement |
| 997531000000108 | Letter received |
| 25064002 | Advice about treatment given |
| 185220009 | Standing height |
| 494131000000105 | Serum alanine aminotransferase level |
| 314940005 | Serum albumin level |
| 167273002 | Standard ECG |
| 397803000 | Syringing ear to remove wax |
| 270424005 | Seen in ophthalmology clinic |
| 164847006 | Advice to patient - subject |
| 171024009 | Seen in neurology clinic |
| 1016851000000100 | Blood test requested |
| 248333004 | Serum potassium level |
| 1321000000100 | Nucleated red blood cell count |
| 185222001 | Body weight |
| 715931000000100 | Serum urea level |
| 1000821000000100 | Serum sodium level |
| 198291000000105 | Serum creatinine level |
| 21522001 | Ca cervix - screening done |
| 1000751000000100 | NHS Health Check invitation first letter |
| 1000731000000100 | Renal profile |
| 1000951000000100 | Serum TSH (thyroid stimulating hormone) level |

| | |
|---|---|
| 27113001 | Serum high density lipoprotein cholesterol level |
| 1000651000000100 | Pulse rate |
| 182531007 | Seen in ENT clinic |
| 185230000 | Serum cholesterol level |
| 1022461000000100 | Serum alkaline phosphatase level |
| 268543007 | Dressing of wound |
| 1000661000000100 | Seen in radiology department |
| 163133003 | Letter from outside agency |
| 185223006 | Serum total bilirubin level |
| 160617001 | Stopped smoking |
| 19231000000106 | Haematocrit |
| 314471005 | Lymphocyte count |
| 160573003 | Haemoglobin estimation |
| 185181002 | Abdomen examined - NAD |
| 1005671000000100 | Patient offered choice of provider |
| 78564009 | MCH - Mean corpuscular haemoglobin |
| 997591000000109 | Blood sample -> Lab NOS |
| 359748005 | Red blood cell count |
| 1022791000000100 | Urine dipstick test |
| 1000621000000100 | Total white cell count |
| 1005681000000100 | Platelet count |
| 1022511000000100 | Seen in general surgery clinic |
| 286261000000101 | Fax sent to: |
| 359748005 | Patient's condition the same |
| 183545006 | MCV - Mean corpuscular volume |
| 1010671000000100 | Neutrophil count |
| 1022291000000100 | Forms - miscellaneous |
| 1022581000000100 | Diet health education |
| 24671000000101 | Telephone call to a patient |
| 1006761000000100 | Plasma glucose level |
| 400010006 | Monocyte count |
| 201101007 | Bone profile |
| 168731009 | Bowel cancer screening programme: faecal occult blood result |
| 408947007 | Eosinophil count |
| 1941000000108 | Serum triglycerides level |
| 82078001 | Declined consent for short message service text messaging |
| 1022551000000100 | Electronic record notes summary verified |

| | |
|---|---|
| 699237001 | Letter sent to patient |
| 14221000000108 | Serum cholesterol/high density lipoprotein ratio |
| 1007881000000100 | Screening - general |
| 70153002 | Hypertension monitoring |
| 1022591000000100 | No known allergy |
| 1022431000000100 | Failed encounter - no answer when rang back |
| 1022651000000100 | Discharge summary report |
| 12063002 | Serum low density lipoprotein cholesterol level |
| 1022491000000100 | X-ray report received |
| 519211000000103 | Basophil count |
| 25581000000101 | Urinalysis = no abnormality |
| 249274008 | Serum calcium level |
| 80313002 | Lloyd George record received |
| 267102003 | Telephone consultation |
| 185276001 | ECG |
| 62315008 | Seen in physiotherapy department |
| 167221003 | Diet good |
| 1015681000000100 | Urine nitrite negative |
| 2051000000104 | Has authorisation for medication under PSD (patient specific direction) |
| 1005691000000100 | Advice given |
| 716186003 | Med3 certificate issued to patient |
| 1022191000000100 | Urine ketone test negative |
| 1022561000000100 | Urea and electrolytes level |
| 14760008 | Lloyd George culled and summarised |
| 11441004 | Refer to physiotherapist |
| 368481000000103 | MCHC - Mean corpuscular haemoglobin concentration |
| 272039006 | Health education - alcohol |
| 185337004 | Standard chest X-ray |
| 15188001 | Letter encounter to patient |
| 49218002 | Haemoglobin A1c level - International Federation of Clinical Chemistry and Laboratory Medicine standardised |
| 275944005 | Letter sent to consultant |
| 935051000000108 | Minor surgery done - injection |
| 167287002 | Urine sample sent to Lab |
| 23056005 | Urine blood test = negative |
| 160639004 | Reassurance given |
| 164226003 | Aerobic exercise 3+ times/week |
| 165339009 | Mole of skin |

| | |
|---|---|
| 314138001 | Seen in gastroenterology clinic |
| 14679004 | High fibre diet |
| 386472008 | Brief intervention for physical activity completed |
| 999791000000106 | ESR - erythrocyte sedimentation rate |
| 84089009 | Seasonal influenza vaccination |
| 47933007 | Inhaler technique - good |
| 171324002 | Wax in ear |
| 1000971000000100 | Review of medication |
| 183039008 | Electrocardiographic monitoring |
| 1746291000006100 | Ex-moderate cigarette smoker (10-19/day) |
| 195967001 | Test result to patient by telephone |
| 185245007 | Health education offered |
| 367391008 | Hormone replacement therapy |
| 15805002 | GPPAQ hours in last week spent walking - 3 hours or more |
| 202855006 | New patient screening done |
| 271739002 | Blood withdrawal for testing |
| 822851000000102 | Patient registration data verified |
| 375031000000109 | New patient screening |
| 170625000 | Occupations |
| 168336001 | Transfer-degraded record entry |
| 49049000 | Aerobic exercise 2 times/week |
| 266717002 | Moderate drinker - 3-6u/day |
| 1022261000000100 | Seen in accident and emergency department |
| 165357005 | Minor surgery done + claimable |
| 24079001 | Serum adjusted calcium concentration |
| 300471006 | GPPAQ hrs in last wk spent in physical exercise - 3hrs or more |
| 60862001 | Electrocardiographic procedure |
| 713019007 | General surgical referral |
| 1746371000006100 | GPPAQ usual level of walking pace - steady |
| 414941008 | Discussion about disorder |
| 195742007 | Cardiac disease monitoring |
| 171175005 | Consultation |
| 196411000000103 | Liquid based cervical cytology screening |
| 183665006 | Indirect encounter |
| 174184006 | Medication given |
| 16991000000107 | Seen by optician |
| 38341003 | GPPAQ not in employment |

| | |
|---|---|
| 366121000000108 | ECG normal |
| 25721000000107 | Alcohol intake |
| 703938007 | O/E - blood pressure reading |
| 183542009 | Spirometry screening |
| 254701007 | Discharged from hospital |
| 394700004 | Scanned document |
| 182833002 | O/E - heart sounds normal |
| 77176002 | General practice physical activity questionnaire physical activity index: inactive |
| 1746271000006100 | Follow-up arranged |
| 185316007 | Raised blood pressure |
| 185719002 | Lifestyle advice regarding hypertension |
| 10601006 | Patient advised about exercise |
| 164854000 | No response to bowel cancer screening programme invitation |
| 223471001 | Chaperone offered |
| 1104081000000100 | Laboratory test requested |
| 443402002 | Acute kidney injury warning stage |
| 24761000000103 | Refer to practice nurse |
| 183616001 | Referral letter |
| 29303009 | Message given to patient |
| 163074001 | Patient's condition worsened |
| 171255006 | Seen by practice phlebotomist |
| 217082002 | Seen in minor injuries department |
| 68566005 | Enjoys heavy exercise |
| 183544005 | Provision of chaperone refused |
| 163020007 | Spirometry |
| 173422009 | Discussion about therapy |
| 276074009 | Choose and book electronic referral letter sent |
| 11429006 | Inhaler technique observed |
| 373251000000108 | Did not attend |
| 401118009 | Diagnostic colonoscopy |
| 183073003 | QRISK2 cardiovascular disease 10 year risk score |
| 268509003 | Failed encounter - message left with household member |
| 49650001 | Seen by consultant |
| 417036008 | E-mail sent to patient |
| 182832007 | Drug compliance checked |
| 993381000000106 | Urine culture |
| 160633003 | Medication review without patient |

| | |
|---|---|
| 239872002 | Urine leukocyte test = negative |
| 92531000000104 | Ex-light cigarette smoker (1-9/day) |
| 25611000000107 | Removal of suture from skin |
| 86094006 | Patient registration |
| 419603000 | Clinical history and observation findings |
| 266922007 | Mail administration procedure |
| 271299001 | Dermatological referral |
| 74506000 | Failed encounter NOS |
| 127783003 | Ambulatory blood pressure recording |
| 763380007 | Serum ferritin level |
| 281399006 | Serum electrolytes level |
| 184047000 | Report administration |
| 1.00064E+15 | Ethnic category not stated - 2001 census |
| 183096003 | Self-help advice leaflet given |
| 307780009 | 12 lead ECG |
| 17741008 | Application of dressing |
| 391156007 | Current smoker |
| 74016001 | No breathlessness |
| 41446000 | Resp. system examined - NAD |
| 394717006 | Low fat diet education |
| 167217005 | Drug compliance good |
| 718087004 | Laboratory test observable |
| 513861000000102 | Framingham coronary heart disease 10 year risk score |
| 113091000000109 | Plasma fasting glucose level |
| 176178006 | History / symptoms |
| 185293001 | ENT referral |
| 183523005 | Histology laboratory test |
| 1000641000000100 | GPPAQ hours in last week spent gardening/DIY - none |
| 170614009 | Liver function tests - general |
| 3895009 | Breast neoplasm screening normal |
| 450759008 | Nursing procedure |
| 183518005 | Letter sent to outside agency |
| 401270003 | Urine test for glucose |
| 408373006 | Trivial drinker - <1u/day |
| 609558009 | Non-smoker |
| 139394000 | A&E report |
| 160237006 | On examination - right dorsalis pedis pulse present |

| | |
|---|---|
| 223485006 | Health education - breast examination |
| 713541000000102 | Seen in vascular clinic |
| 1003181000000100 | Alcohol consumption |
| 185190009 | Blood test due |
| 75088002 | Urine examination |
| 161938003 | Examination / signs |
| 160593006 | MED5 issued to patient |
| 993411000000108 | Lloyd George and problem summary |
| 185438005 | On examination - left dorsalis pedis pulse present |
| 275926002 | Aerobic exercise 1 time/week |
| 1371000000101 | Serum folate level |
| 78809005 | Medication review done by pharmacist |
| 992831000000108 | Consent given for blood test |
| 160637002 | Prescription collected by pharmacy |
| 125605004 | Seen in audiology clinic |
| 44054006 | Minor surgery done - excision |
| 9632001 | Email sent to patient |
| 9826008 | O/E - gait normal |
| 184103008 | Clinical letter |
| 182884001 | Travel destination |
| 363746003 | Serum total protein |
| 823691000000103 | Communication from: |
| 417662000 | eConsultation via online application |
| 719329004 | DNA hospital appointment |
| 24561000000109 | Vascular disease risk assessment |
| 939511000000101 | Administration note |
| 23919004 | Registered for online access to local practice |
| 268926000 | Bereavement |
| 177860007 | Motor vehicle traffic accident |
| 314481009 | Alcohol intake within recommended sensible limits |
| 183061003 | Screening procedure |
| 397881000 | Patient medical record envelope received from family practitioner committee |
| 10509002 | Light drinker - 1-2u/day |
| 77477000 | Incoming mail NOS |
| 162356005 | Diagnostic cystoscopy |
| 161894002 | Consent given to share patient data with specified third party |
| 16791000000109 | Breast neoplasm screening |

| | |
|---|---|
| 195647007 | Over the counter aspirin therapy |
| 315217005 | Dissent from disclosure of personal confidential data by Health and Social Care Information Centre |
| 235595009 | Dissent from secondary use of general practitioner patient identifiable data |
| 34014006 | PSA (prostate-specific antigen) level |
| 1031081000000100 | Seen in walk in centre |
| 1746451000006100 | Brief intervention for excessive alcohol consumption completed |
| 163120009 | Chest clear |
| 198436008 | Histopathology test |
| 165349007 | Normal histology findings |
| 8392000 | Test request : Full Blood Count |
| 170638008 | Last menstrual period -1st day |
| 268547008 | Diagnostic arthroscopy of knee joint |
| 163684001 | Gastroscopy |
| 17293009 | Letter invite to screening |
| 313148005 | Medication changed |
| 440401008 | Smoking cessation education |
| 364712009 | Patient reassurance |
| 261000000103 | Medication management |
| 37351000000107 | Seen by specialist physician |
| 394726009 | Cervical smear - 1st call |
| 170635006 | Seen in hospital out-pat. |
| 183521007 | Intramuscular injection |
| 1030791000000100 | Influenza vaccination invitation first letter sent |
| 267032009 | Gynaecological referral |
| 1746411000006100 | Urine protein test = + |
| 343121000000108 | Ex-heavy cigarette smoker (20-39/day) |
| 185248009 | Tobacco smoking consumption |
| 705055006 | Computer summary updated |
| 308021002 | ENT examination - NAD |
| 40701008 | 10g monofilament sensation R foot normal |
| 30037006 | Adult health examination |
| 306635009 | Referral to G.P. |
| 287721000000104 | Seen in out of hours centre |
| 522261000000101 | Low cholesterol diet education |
| 203351000000102 | Drugs not issued |
| 183830008 | Minor surgery done - cautery |

| | |
|---|---|
| 47382004 | Prescription collected by patient |
| 281794004 | On examination - right posterior tibial pulse present |
| 182918009 | Fall - accidental |
| 394670005 | INR - international normalised ratio |
| 90834002 | BP screening - first recall |
| 17369002 | Referral letter sent |
| 105542008 | Choose and book referral |
| 161920001 | New registration check done and claimable |
| 267432004 | Total cholesterol:HDL (high density lipoprotein) ratio |
| 24420007 | Cardiovascular disease risk assessment done |
| 167297006 | Adult screening |
| 1022481000000100 | Orthopaedic referral |
| 310500000 | SMS text message received from patient |
| 302415002 | GPPAQ hours in last week spent cycling - none |
| 270224008 | Married |
| 53726008 | Serum free T4 level |
| 14361000000109 | Incoming mail |
| 35489007 | Consent given for communication by text messaging |
| 171302002 | Microbiology |
| 1016971000000100 | MSU sent for C/S |
| **After the confirmation of PD diagnosis** | |
| **SNOMED codes** | **Description** |
| 279991000000102 | SMS (short message service) text message sent to patient |
| 428481002 | Patient mobile telephone number |
| 185230000 | Seen in neurology clinic |
| 313334002 | Blood sample taken |
| 185317003 | Telephone encounter |
| 182836005 | Telephone consultation |
| 25721000000107 | Never smoked tobacco |
| 266919005 | Template entry - EHR composition type |
| 182771004 | Medication review |
| 386472008 | Informed consent for procedure |
| 88551000000109 | Medication review with patient |
| 788007007 | General practice service |
| 314530002 | Medication review done |
| 162999005 | O/E - pulse rhythm regular |
| 310080006 | Pharmacy service |

| | |
|---|---|
| 270426007 | Did not attend - no reason |
| 184103008 | Weight monitoring |
| 270426007 | Patient telephone number |
| 307818003 | Immunisation course to maintain protection against SARS-CoV-2 (severe acute respiratory syndrome coronavirus 2) |
| 1362591000000100 | Medication requested |
| 182888003 | Administrative procedure |
| 14734007 | Ex-smoker |
| 8517006 | Failed encounter - message left on answer machine |
| 185324002 | Failed encounter |
| 185340004 | Administration note |
| 37351000000107 | Preferred method of contact: unknown |
| 822701000000109 | Has authorisation for medication under PSD (patient specific direction) |
| 1990681000006100 | eConsultation via online application |
| 1068881000000100 | Enhanced review indicated before granting access to own health record |
| 185210004 | Choice and booking enhanced services administration |
| 1364731000000100 | Systolic arterial pressure |
| 72313002 | Diastolic arterial pressure |
| 222711000000102 | GFR (glomerular filtration rate) calculated by abbreviated Modification of Diet in Renal Disease Study Group calculation |
| 1091811000000100 | Letter from specialist |
| 1020291000000100 | Telephone triage encounter |
| 270425006 | Serum potassium level |
| 401267002 | Serum urea level |
| 1000651000000100 | Informing patient of named accountable general practitioner |
| 1000731000000100 | Serum sodium level |
| 1000661000000100 | Serum creatinine level |
| 908481000000105 | FBC - full blood count |
| 1000951000000100 | Body weight |
| 1022441000000100 | Patient allocated named accountable general practitioner |
| 11816003 | Serum alanine aminotransferase level |
| 27113001 | Haemoglobin estimation |
| 965831000000103 | Serum alkaline phosphatase level |
| 1018251000000100 | Letter from consultant |
| 1022431000000100 | Urea and electrolytes level |
| 1000971000000100 | Platelet count |
| 1000621000000100 | MCV - Mean corpuscular volume |
| 183049006 | Red blood cell count |

| | |
|---|---|
| 275693003 | Patient given advice |
| 394617004 | Monocyte count |
| 1022581000000100 | Neutrophil count |
| 1000821000000100 | Seen in hospital casualty |
| 1022651000000100 | Result |
| 1022571000000100 | Lymphocyte count |
| 1022451000000100 | Haematocrit |
| 1022491000000100 | Total white cell count |
| 1022551000000100 | Signposting |
| 975131000000104 | Eosinophil count |
| 699237001 | Serum albumin level |
| 1022591000000100 | Basophil count |
| 1022561000000100 | Bowel cancer screening programme faecal occult blood test normal |
| 1022291000000100 | Discharge summary |
| 1022541000000100 | Medication review of medical notes |
| 182884001 | Pulse rate |
| 78564009 | Bowel cancer screening programme: faecal occult blood result |
| 375211000000108 | MCH - Mean corpuscular haemoglobin |
| 368481000000103 | Drug compliance good |
| 1022471000000100 | Seen by practice phlebotomist |
| 93311000000106 | Haemoglobin A1c level - International Federation of Clinical Chemistry and Laboratory Medicine standardised |
| 373942005 | Patient contact administration |
| 185305000 | Standing height |
| 999791000000106 | Blood pressure recorded by patient at home |
| 401271004 | Protocol entry |
| 714241000000102 | Red blood cell distribution width |
| 413153004 | Body mass index |
| 394837003 | Administrative reason for encounter |
| 185351004 | Seen in ophthalmology clinic |
| 248333004 | Letter encounter |
| 60621009 | Patient review |
| 993501000000105 | Telephone call to a patient |
| 308720009 | SMS text message sent to patient |
| 185222001 | E-mail sent to patient |
| 763288003 | Seen in orthopaedic clinic |
| 24671000000101 | White British - ethnic category 2001 census |
| 494131000000105 | NHS 111 report received |

| | |
|---|---|
| 26079004 | Serum calcium level |
| 887641000000105 | MCHC - Mean corpuscular haemoglobin concentration |
| 304507003 | Serum total bilirubin level |
| 1000691000000100 | Serum total protein |
| 907151000000108 | Serum cholesterol level |
| 997591000000109 | Letter received |
| 1005671000000100 | Serum triglycerides level |
| 1022481000000100 | Long term condition care planning invitation first letter |
| 1000811000000100 | Serum lipids level |
| 198291000000105 | Serum high density lipoprotein cholesterol level |
| 912681000000107 | Serum TSH (thyroid stimulating hormone) level |
| 1005661000000100 | Seen in dermatology clinic |
| 1022791000000100 | Brief history taken |
| 1005681000000100 | Administration |
| 408363009 | Patient understands why taking all medication |
| 270422009 | Serum low density lipoprotein cholesterol level |
| 160240006 | Informed consent given |
| 225323000 | Serum cholesterol/high density lipoprotein ratio |
| 1015681000000100 | Plan |
| 1022191000000100 | Enjoys light exercise |
| 160631001 | Lifestyle education |
| 203031000000107 | Moderate frailty |
| 717371000000107 | Serum adjusted calcium concentration |
| 955691000000108 | Seasonal influenza vaccination given by pharmacist |
| 313204009 | Mail administration procedure |
| 935051000000108 | Clinical letter |
| 185276001 | Seen in physiotherapy department |
| 976631000000101 | Serum folate level |
| 416118004 | Consent given for blood test |
| 993381000000106 | White: English or Welsh or Scottish or Northern Irish or British - England and Wales ethnic category 2011 census |
| 993411000000108 | Thyroid function test |
| 59621000 | Standard ECG |
| 907131000000101 | Urine nitrite negative |
| 314138001 | History/symptoms |
| 308021002 | Serum ferritin level |
| 11429006 | Consultation |
| 1364181000000100 | Direct-acting oral anticoagulant dose unchanged |

| | |
|---|---|
| 164847006 | Implied consent for core Summary Care Record dataset upload |
| 1016851000000100 | NHS Health Check completed |
| 160237006 | Blood test requested |
| 307780009 | Health education - exercise |
| 167297006 | Date |
| 167261002 | Urine blood test = negative |
| 773011000000101 | Urine glucose test negative |
| 993391000000108 | Did not attend |
| 413672003 | Serum vitamin B12 level |
| 279039007 | Liver function test |
| 25781000000108 | Urinalysis = no abnormality |
| 523221000000100 | Has shown no side effects from medication |
| 1001231000000100 | Awaiting clinical code migration to EMIS Web |
| 49218002 | Influenza vaccination invitation first short message service text message sent |
| 314940005 | Review of medication |
| 777791000000102 | Consent given for communication by text messaging |
| 281399006 | Serum globulin level |
| 1572871000006100 | Seen in ENT clinic |
| 408508002 | Seen in clinic |
| 167221003 | SARS-CoV-2 (severe acute respiratory syndrome coronavirus 2) vaccination invitation SMS (short message service) text message sent |
| 1240781000000100 | Audit administration |
| 185223006 | Seasonal influenza vaccination given by other healthcare provider |
| 705025004 | Consent given for communication by short message service text messaging |
| 105542008 | Mammography normal |
| 250171008 | Clinical history and observation findings |
| 168749009 | Consent given for communication by email |
| 713481000000109 | Percentage basophils |
| 955651000000100 | Out of hours report |
| 165355002 | Urine albumin:creatinine ratio |
| 394700004 | Face to face consultation |
| 1015501000000100 | Patient identity verified |
| 439811001 | Notes summary on computer |
| 8392000 | Urine protein test negative |
| 736728004 | Seen by ambulance crew |
| 170557005 | British or mixed British - ethnic category 2001 census |

| | |
|---|---|
| 1023491000000100 | Non-smoker |
| 430193006 | Chronic disease initial assessment |
| 184229000 | Patient informed - test result |
| 306806004 | Smoking cessation advice |
| 167273002 | Diet health education |
| 183616001 | Main spoken language English |
| 185228002 | Urine ketone test negative |
| 719329004 | High risk category for developing complication from COVID-19 infection |
| 167287002 | Admin. reminder |
| 713776001 | Urine creatinine level |
| 1875551000006100 | Medication review done by pharmacist |
| 1007881000000100 | Urine dipstick test |
| 1300561000000100 | Follow-up arranged |
| 911361000000104 | Consent given to receive test results by short message service text messaging |
| 1003271000000100 | Repeat medication check |
| 763380007 | Hypertension monitoring |
| 315570003 | GPPAQ hours in last week spent cycling - none |
| 182838006 | Alcohol intake |
| 29857009 | Seen in urology clinic |
| 773051000000102 | Express consent for core and additional Summary Care Record dataset upload |
| 286261000000101 | Seen in gastroenterology clinic |
| 49727002 | Serum non high density lipoprotein cholesterol level |
| 185184005 | Urine sample sent to Lab |
| 185381007 | 12 lead ECG |
| 1746291000006100 | Seen in speech and language clinic |
| 268400002 | Peripheral oxygen saturation |
| 431314004 | Test result to patient by telephone |
| 185220009 | Diet education |
| 275944005 | Serum PSA (prostate specific antigen) level |
| 165339009 | Diet good |
| 160573003 | Letter sent to patient |
| 185337004 | Random sample |
| 185293001 | Magnetic resonance imaging |
| 308447003 | At risk of dementia |
| 113091000 | General practice physical activity questionnaire physical activity index: active |

| | |
|---|---|
| 165357005 | Implementation of aseptic technique |
| 1127441000000100 | Seen by consultant |
| 840671000000108 | Prescription |
| 370822003 | Medication review done by clinical pharmacist |
| 366241000000106 | Seen in cardiac clinic |
| 260885003 | GPPAQ not in employment |
| 310500000 | Failed encounter - no answer when rang back |
| 50417007 | Medicines reconciliation completed |
| 14760008 | Consent given to share patient data with specified third party |
| 1746171000006100 | Referral to social prescribing service |
| 1000381000000100 | Referral letter sent |
| 698464007 | Email sent to outside agency |
| 46825001 | Seen in GP's surgery |
| 170625000 | Inhaler technique - good |
| 183093006 | Mild frailty |
| 185321005 | Administration of medication under patient specific direction |
| 928971000000108 | Provision of chaperone refused |
| 195967001 | Letter encounter to patient |
| 715931000000100 | Discharged from hospital |
| 709901000000106 | Enjoys moderate exercise |
| 319951000000105 | Medication review without patient |
| 185202000 | Constipation |
| 408562003 | Urine microalbumin level |
| 925791000000100 | QOF (Quality and Outcomes Framework) quality indicator-related care invitation |
| 160632008 | Telephone call from a patient |
| 413973005 | Breast neoplasm screening normal |
| 170638008 | Urine leucocyte test = negative |
| 526421000000109 | Blood pressure monitoring invitation SMS (short message service) text message |
| 183665006 | Indication for each drug checked |
| 1010251000000100 | New medication commenced |
| 35489007 | Repeat prescription reviewed by pharmacist |
| 968191000000100 | Interpreter not needed |
| 401176004 | Patient contact details verified |
| 1022511000000100 | Seen in radiology department |
| 1085181000000100 | Neurology |
| 394591006 | Estimated creatinine clearance (Cockcroft-Gault formula) |

| | |
|---|---|
| 408343002 | Scanned document |
| 811921000000103 | Physical examination |
| 1109921000000100 | Long term condition care planning invitation second letter |
| 903081000000107 | Medication Reconciliation |
| 266712008 | Third party encounter |
| 185181002 | Seen by neurologist |
| 183452005 | Seen in general medical clinic |
| 161891005 | Informed consent given for treatment |
| 315595002 | Seen in out of hours centre |
| 45326000 | Drug side effects checked |
| 912721000000100 | NHS Health Check invitation short message service text message |
| 5880005 | Serum bicarbonate level |
| 408947007 | Discharge summary report |
| 370208006 | Marital state unknown |
| 95324001 | Hospital prescription |
| 171024009 | Medication changed |
| 305684006 | Advice about treatment given |
| 1001371000000100 | Serum chloride level |
| 185318008 | No new symptoms |
| 762911000000102 | Non - drinker |
| 980641000000103 | Patient's next of kin |
| 1957991000006100 | Influenza vaccination invitation first letter sent |
| 170635006 | Sampling of cervix for Papanicolaou smear done |
| 24681000000104 | Had a chat to patient |
| 408356009 | Emergency hospital admission |
| 185241003 | Referral letter sent by email |
| 928451000000107 | Serum CRP (C reactive protein) level |
| 396275006 | NHS Health Check invitation |
| 200481000000107 | Seen in musculoskeletal clinic |
| 201101007 | Dietary advice |
| 1000681000000100 | ESR - erythrocyte sedimentation rate |
| 21522001 | Able to use private transport |
| 523241000000107 | Lloyd George record received |
| 394995008 | Administration of medication under patient group direction |
| 314503007 | Post hospital discharge medication reconciliation with medical notes |
| 928991000000107 | Serum bilirubin level |
| 1000671000000100 | Alert received from telehealth monitoring system |

| | |
|---|---|
| 30989003 | Medication management |
| 398838000 | Administration of first dose of SARS-CoV-2 (severe acute respiratory syndrome coronavirus 2) vaccine |
| 1746501000006100 | Pulse regular |
| 401270003 | Health education - alcohol |
| 711124002 | Administration of second dose of SARS-CoV-2 (severe acute respiratory syndrome coronavirus 2) vaccine |
| 427933004 | GPPAQ usual level of walking pace - slow |
| 999691000000104 | Seen by clinical pharmacist |
| 307541003 | Initial memory assessment |
| 182832007 | No breathlessness |
| 165791000000107 | Patient advised about exercise |
| 492811000000103 | General practice physical activity questionnaire physical activity index: moderately inactive |
| 1127431000000100 | Blood test due |
| 1324691000000100 | ECG |
| 404640003 | Advice given |
| 730061000000107 | Reason for referral |
| 14361000000109 | Phlebotomy generated from secondary care done by practice |
| 183073003 | Infection screening |
| 243790003 | Bone profile |
| 888901000000102 | Provision of patient satisfaction questionnaire |
| 271636001 | Seen in mental health clinic |
| 300627003 | Seen in elderly care clinic |
| 185719002 | Serum free T4 level |
| 1324681000000100 | In-house physio |
| 864231000000108 | Referred for health coaching |
| 699249000 | Letter encounter from patient |
| 335081000000105 | Triage |
| 1006761000000100 | Additional note |
| 418043000 | Education about alcohol consumption |
| 431957001 | Temperature |
| 77248004 | Medication review done by pharmacy technician |
| 185248009 | Good compliance with inhaler |
| 718087004 | Wound care |
| 716101000000104 | Seen by physiotherapist |
| 182878004 | Repeat prescription monitoring |
| 160629005 | Medication review invitation |

| | |
|---|---|
| 399208008 | Current non-smoker |
| 165333005 | Home visit |
| 276221000000100 | Infection control procedure |
| 719326006 | Seen in colorectal clinic |
| 270424005 | GPPAQ hours in last week spent walking - 3 hours or more |
| 167223000 | General practice physical activity questionnaire physical activity index: inactive |
| 1746271000006100 | Referral for further care |
| 4525004 | General practice physical activity questionnaire physical activity index: moderately active |
| 408373006 | Medication increased |
| 310065000 | Referral to physiotherapist |
| 390862006 | X-ray report received |
| 366979004 | Advice to continue with drug treatment |
| 855981000006102 | Medication monitoring |
| 281078001 | Drug compliance checked |
| 23056005 | Telehealth monitoring |
| 736601004 | Non-diabetic hyperglycaemia |
| 1000751000000100 | Open access service |
| 185285001 | QRISK2 cardiovascular disease 10 year risk score |
| 271739002 | ECG: sinus rhythm |
| 366121000000108 | Renal profile |
| 183444007 | Seen in pain clinic |
| 394671009 | Demonstration of inhaler technique |
| 37361000000105 | Seen by occupational therapist |
| 38341003 | Phacoemulsification of lens and insertion of prosthetic replacement |
| 2071000000108 | Appointment made |
| 225358003 | Letter from Driver and Vehicle Licensing Authority received |
| 703421000 | Seen in general surgery clinic |
| 170922004 | GPPAQ hrs in last wk spent in physical exercise - 3hrs or more |
| 295051000000101 | Exercise physically impossible |
| 401250009 | Exercise education |
| 426783006 | Seen in accident and emergency department |
| 1087551000000100 | Short message service text message sent |
| 440168007 | Patient given telephone advice out of hours |
| 700449008 | Has authorisation for medication under PGD (patient group direction) |
| 160618006 | Provision of advice, assessment or treatment limited due to COVID-19 pandemic |

| | |
|---|---|
| 711408005 | Synchronisation of repeat medication |
| 185287009 | Advice to return if problem persists or deteriorates |
| 313184003 | Self-help advice leaflet given |
| 1672851000006100 | Invitation to participate in research study |
| 172630005 | Intramuscular injection of vitamin B12 |
| 185244006 | 10g monofilament sensation R foot normal |
| 394675000 | No response to bowel cancer screening programme invitation |
| 439708006 | Liquid based cervical cytology screening |
| 1746371000006100 | Opportunistic verification of patient mobile telephone number |
| 395067002 | White British |
| 249274008 | ECG normal |
| 314373007 | Smoking monitoring invitation |
| 197480006 | Wheeze absent |
| 713056003 | Discussion about clinical red flag warning sign |
| 315236000 | Depression screening using questions |
| 810901000000102 | Plasma glucose level |
| 170614009 | Optimisation of drug dosage |
| 735259005 | Drugs not issued |
| 185731000 | Failed encounter - short message service text message delivery failure |
| 185229005 | Reassurance given |
| 394712000 | Inhaler technique observed |
| 927621000000101 | No cough |
| 822721000000100 | GPPAQ hrs in last wk spent gardening/DIY-some but less than 1hr |
| 1746461000006100 | Email received from third party |
| 1010671000000100 | 10g monofilament sensation L foot normal |
| 310436000 | Nucleated red blood cell count |
| 417036008 | Seen by Accident and Emergency service |
| 77477000 | CHA2DS2-VASc (congestive heart failure, hypertension, age 2, diabetes mellitus, stroke 2, vascular disease, age, sex category) stroke risk score |
| 415693003 | SMS text message received from patient |
| 184156005 | Email encounter from third party |
| 82078001 | Urine leucocyte test = + |
| 134395001 | 360 degree sweep of cervix performed |
| 183096003 | Urinary symptoms |
| 183039008 | Provision of copy of letter from specialist to patient |
| 1321171000000100 | Has a carer |
| 68566005 | Dressing of wound |
| 373251000000108 | Incoming mail |

| | |
|---|---|
| 394703002 | Magnetic resonance imaging of spine |
| 182850000 | Vitamin D deficiency |
| 515201000000103 | O/E - Left posterior tibial pulse normal |
| 1066951000000100 | CT of head |
| 182531007 | Syringing ear to remove wax |
| 32485007 | Patient feels well |
| 310429001 | Outcome |
| 185227007 | Seen by Primary Care Navigator |
| 394669009 | Respiratory rate |
| 162415008 | O/E - right posterior tibial pulse normal |
| 200971000000100 | Consent given for minor surgery procedure |
| 185307008 | Fax sent to: |
| 161898004 | Cancer care review |
| 720006006 | O/E - right dorsalis pedis normal |
| 1052411000000100 | NHS Health Check verbal invitation |
| 163133003 | GPPAQ usual level of walking pace - steady |
| 811611000000101 | Abdomen examined - NAD |
| 1746511000006100 | Chest clear |
| 34713006 | Electronic record notes summary verified |
| 414907000 | Medical records review |
| 1746431000006100 | Neurological referral |
| 10601006 | GPPAQ hours in last week spent in physical exercise - none |
| 802241000000106 | Community health services |
| 183631005 | Blood sample -> Lab NOS |
| 241645008 | No known allergy |
| 717661000000106 | Seen in hospital out-pat. |
| 170777000 | Alcohol units consumed per week |
| 267112005 | Brief intervention for physical activity completed |
| 198241000000100 | Wax in ear |
| 271921002 | O/E - left dorsalis pedis normal |
| 308722001 | Smoking cessation education |
| 14221000000108 | Nocturia |
| 134420004 | Light drinker - 1-2u/day |
| 185271006 | No suicidal thoughts |
| 15188001 | Provision of patient questionnaire |
| 871691000000100 | Planned telephone contact |
| 335661000000109 | Complaining of a general symptom |

| | |
|---|---|
| 522261000000101 | Short message service text message received from patient |
| 414912004 | Self measured blood pressure reading |
| 301708006 | Social prescribing offered |
| 160575005 | Treatment plan given |
| 148561000000109 | Medicines adherence checked |
| 314705003 | GPPAQ hrs in last wk spent on house work/child care-3hrs or more |
| 162409008 | E-mail received from patient |
| 413294000 | Structured medication review |
| 312853008 | Colonoscopy |
| 303653007 | Advice given about weight management |
| 168731009 | ECG finding |
| 1746241000006100 | Medication review due |
| 428489000 | Inhaler technique not checked |
| 86290005 | Seen in breast clinic |
| 1082641000000100 | Signposting to GP (general practitioner) |
| 1239511000000100 | Falls |
| 190634004 | QOF (Quality and Outcomes Framework) non-diabetic hyperglycaemia quality indicator-related care invitation |
| 716186003 | Seen in respiratory clinic |
| 314529007 | Message given to patient |
| 1084071000000100 | English - ethnic category 2001 census |
| 526431000000106 | Referral to ear, nose and throat service |
| 139394000 | Clotting screening test |
| 185211000 | Seen in audiology clinic |
| 414896001 | Seen by general practitioner |
| 81680005 | Removal of skin sutures/clips |
| 698471002 | GPPAQ hours in last week spent gardening/DIY - none |
| 267039000 | Tobacco smoking behaviour - finding |
| 162290004 | GPPAQ hrs last wk spent house work/child care-1hr less than 3hrs |
| 200491000000109 | Influenza vaccination invitation second letter sent |
| 1011481000000100 | Prothrombin time |
| 852471000000107 | NHS Health Check invitation first letter |
| 185190009 | Teetotaller |
| 254701007 | Using inhaled steroids - normal dose |
| 84229001 | Chaperone present |
| 16310003 | Patient's condition improved |
| 239873007 | Bowel cancer screening programme |

**Supplementary Table 5. Codes used to search for each of the top 10 most common co-existing diagnoses before and after the confirmation of Parkinson's disease.**

| Before the confirmation of PD diagnosis | | |
|---|---|---|
| **Diagnosis** | **SnomedCTConceptId** | **Term** |
| **Musculoskeletal and connective tissue diseases** | 81680005 | Neck pain |
| | 279039007 | Low back pain |
| | 45326000 | Shoulder pain |
| | 49218002 | Hip pain |
| | 30989003 | Knee pain |
| | 47933007 | Foot pain |
| | 239873007 | Osteoarthritis of knee |
| | 279069000 | Musculoskeletal pain |
| | 202794004 | Lumbago with sciatica |
| | 281245003 | Musculoskeletal chest pain |
| | 201819000 | Generalised osteoarthritis |
| | 23056005 | Sciatica |
| | 247366003 | Acute back pain with sciatica |
| | 57676002 | Joint pain |
| | 33952002 | Localised osteoarthritis, unspecified, of the lower leg |
| | 8847002 | Osteoarthritis of spine |
| | 161894002 | C/O - low back pain |
| | 396275006 | Osteoarthritis NOS |
| | 30989003 | Knee joint pain |
| | 427933004 | Seen in musculoskeletal clinic |
| | 239873007 | Osteoarthritis NOS, of knee |
| | 312225001 | Musculoskeletal and connective tissue diseases |
| | 239872002 | Osteoarthritis of hip |
| | 396275006 | Osteoarthritis |
| | 3723001 | Arthritis |
| | 312225001 | Musculoskeletal and connective tissue disorder |
| | 239733006 | Anterior knee pain |
| | 202482009 | Wrist joint pain |
| | 84869007 | Musculoskeletal symptom |
| | 413159000 | Referral to musculoskeletal clinic |
| | 279040009 | Mechanical low back pain |

| | | |
|---|---|---|
| | 239872002 | Osteoarthritis NOS, of hip |
| | 396275006 | Osteoarthritis and allied disorders |
| | 275554004 | [V]Personal history of arthritis |
| | 161567008 | H/O: rheumatoid arthritis |
| | 202480001 | Elbow joint pain |
| | 450521003 | Patellofemoral osteoarthritis |
| | 202490009 | Ankle joint pain |
| | 396275006 | Osteoarthritis NOS, of the lower leg |
| | 268054009 | Osteoarthritis of multiple joints |
| **Skin lesions & dermatitis** | 162415008 | C/O: a rash |
| | 398838000 | Seborrhoeic keratosis |
| | 398838000 | Seborrhoeic wart |
| | 703938007 | Inflammatory dermatosis |
| | 43116000 | Eczema |
| | 201091002 | Skin tag |
| | 271807003 | [D]Rash and other nonspecific skin eruption NOS |
| | 442279002 | Rash of genitalia |
| | 24079001 | Atopic dermatitis/eczema |
| | 304386008 | O/E - itchy rash |
| | 201101007 | Actinic keratosis |
| | 254701007 | Basal cell carcinoma of skin |
| | 268911002 | O/E - rash present |
| | 57092006 | Flexural eczema |
| | 161561009 | H/O: eczema |
| | 40275004 | Contact dermatitis and other eczemas |
| | 80659006 | Disorder of skin and/or subcutaneous tissue |
| | 50563003 | Seborrhoeic dermatitis |
| | 366362000 | Varicose eczema |
| | 80659006 | Disorder of skin AND/OR subcutaneous tissue |
| | 24079001 | Dermatitis/eczemas |
| | 275700003 | Varicose eczema - leg |
| | 40275004 | Contact dermatitis |
| | 445111008 | Cryotherapy of actinic keratosis |
| | 162415008 | Complaining of a rash |

| | | |
|---|---|---|
| | 24079001 | Atopic dermatitis |
| **Hypertensive disorder** | 59621000 | Essential hypertension |
| | 401118009 | Hypertension annual review |
| | 275944005 | Hypertension monitoring |
| | 401048005 | Hypertension six month review |
| | 38341003 | Hypertensive disease |
| | 185723005 | Hypertension monitoring telephone invite |
| | 38341003 | BP - hypertensive disease |
| | 185719002 | Hypertension monitoring first letter |
| | 185720008 | Hypertension monitoring second letter |
| | 308116003 | Antihypertensive therapy |
| | 170586008 | Treatment for hypertension started |
| | 38341003 | Hypertensive disorder |
| | 161501007 | H/O: hypertension |
| | 473225006 | Hypertension medication review |
| | 59621000 | Essential hypertension NOS |
| | 407567007 | Patient on maximal tolerated antihypertensive therapy |
| | 38341003 | Hypertension |
| | 185264001 | Seen in hypertension clinic |
| **Headache & Migraine** | 398057008 | Tension headache |
| | 25064002 | Headache |
| | 37796009 | Migraine |
| | 272027003 | Complaining of a headache |
| | 267096005 | Frontal headache |
| | 161481007 | H/O: migraine |
| | 272027003 | C/O - a headache |
| | 23186000 | Menstrual migraine |
| | 25064002 | [D]Headache |
| | 4969004 | Sinus headache |
| | 193030005 | Migraine variants |
| **Depression & Anxiety** | 413169006 | On depression register |
| | 197480006 | Anxiety disorder |
| | 231504006 | Mixed anxiety and depressive disorder |
| | 198288003 | Anxiety state |

| | | |
|---|---|---|
| | 231504006 | [X]Other mixed anxiety disorders |
| | 35489007 | Depression |
| | 413973005 | Depression interim review |
| | 87414006 | Reactive depression |
| | 58703003 | Postpartum depression |
| | 161469008 | H/O: depression |
| | 35489007 | Depressive disorder |
| | 161470009 | H/O: anxiety state |
| | 207363009 | Anxiety neurosis |
| | 413974004 | Depression medication review |
| | 310495003 | Mild depression |
| | 268621008 | Recurrent major depressive episodes |
| | 87414006 | Reactive depression (situational) |
| | 191616006 | Recurrent depression |
| | 21897009 | Generalised anxiety disorder |
| | 310497006 | Severe depression |
| | 310496002 | Moderate depression |
| **Haemorrhoids & Constipation** | 70153002 | Haemorrhoids |
| | 70153002 | Haemorrhoids NOS |
| | 195453000 | Internal haemorrhoids, simple |
| | 70153002 | Piles - haemorrhoids |
| | 26373009 | External thrombosed haemorrhoids |
| | 14760008 | Constipation NOS |
| | 14760008 | Constipation |
| | 174359003 | Haemorrhoids: ligate/excise |
| **Abdominal Hernia** | 177860007 | Primary repair of inguinal hernia |
| | 84089009 | Hiatus hernia |
| | 396347007 | Umbilical hernia |
| | 84089009 | Hiatal hernia |
| | 413146008 | Primary laparoscopic repair of inguinal hernia |
| | 309752000 | Hiatus hernia with gangrene |
| | 39839004 | Diaphragmatic hernia |
| | 44946007 | Repair of umbilical hernia |
| | 396232000 | Inguinal hernia |
| | 73147001 | Direct inguinal hernia |
| | 236022004 | Left inguinal hernia |

| | | |
|---|---|---|
| | 65626001 | Indirect inguinal hernia |
| | 236021006 | Right inguinal hernia |
| | 1171000119108 | Unilateral recurrent inguinal hernia unspecified |
| | 177862004 | Primary mesh repair of inguinal hernia |
| | 177860007 | Primary repair of inguinal hernia NOS |
| **Respiratory tract infection** | 50417007 | Lower respiratory tract infection |
| | 281794004 | Viral upper respiratory tract infection |
| | 195742007 | Acute lower respiratory tract infection |
| | 275498002 | Respiratory tract infection |
| **Dyspepsia/Indigestion/Reflux** | 162031009 | Dyspepsia |
| | 162031009 | Indigestion symptoms |
| | 266433003 | Reflux oesophagitis |
| | 162031009 | Indigestion |
| | 235595009 | Gastro-oesophageal reflux |
| | 225587003 | Gastric reflux |
| | 249511005 | Flatulent dyspepsia |
| | 266435005 | Gastro-oesophageal reflux disease without oesophagitis |
| | 698065002 | Acid reflux |
| | 414581006 | Laryngopharyngeal reflux |
| | 235595009 | Gastroesophageal reflux disease |
| | 266433003 | Gastro-oesophageal reflux disease with oesophagitis |
| **UTI** | 314940005 | Suspected UTI (urinary tract infection) |
| | 68566005 | Urinary tract infection, site not specified |
| | 197927001 | Recurrent urinary tract infection |
| | 68566005 | Urinary tract infection |
| | 68566005 | Urinary tract infection, site not specified NOS |
| | 68566005 | UTI - Urinary tract infection |
| **After the confirmation of PD diagnosis** | | |
| **Disease** | **SnomedCTConceptId** | **Term** |
| **Musculoskeletal disorders** | 23056005 | Sciatica |
| | 30989003 | Knee pain |

| | | |
|---|---|---|
| | 279039007 | Low back pain |
| | 239872002 | Osteoarthritis of hip |
| | 134407002 | Chronic back pain |
| | 45326000 | Shoulder pain |
| | 396275006 | Osteoarthritis |
| | 3723001 | Arthritis |
| | 323291000119108 | Osteoarthritis of left hip joint |
| | 49218002 | Hip pain |
| | 161891005 | Backache |
| | 202794004 | Lumbago with sciatica |
| | 413159000 | Referral to musculoskeletal clinic |
| | 81680005 | Neck pain |
| | 239873007 | Osteoarthritis of knee |
| | 239874001 | Osteoarthritis of ankle |
| | 427933004 | Seen in musculoskeletal clinic |
| | 279069000 | Musculoskeletal pain |
| | 161894002 | C/O - low back pain |
| | 161891005 | Back pain without radiation NOS |
| | 69896004 | Rheumatoid arthritis |
| | 847261000000104 | Rheumatoid arthritis annual review |
| | 239792003 | Seronegative rheumatoid arthritis |
| | 230821000000102 | Rehabilitation for musculoskeletal disorders |
| | 279035001 | Acute thoracic back pain |
| | 312225001 | Musculoskeletal and connective tissue diseases |
| | 312225001 | Musculoskeletal and connective tissue disorder |
| | 373623009 | Osteoarthritis of glenohumeral joint |
| | 161891005 | Back pain |
| | 239872002 | Osteoarthritis NOS, of hip |
| | 161568003 | H/O: osteoarthritis |
| | 8847002 | Osteoarthritis of spine |
| | 841261000000105 | Referral to community musculoskeletal service |
| | 247366003 | Acute back pain with sciatica |
| | 1.07895E+15 | Referral to musculoskeletal extended scope practitioner |
| | 161894002 | Complaining of low back pain |

| | | |
|---|---|---|
| | 371081002 | Arthritis of knee |
| | 239733006 | Anterior knee pain |
| | 374471000000101 | Referral to musculoskeletal special interest general practitioner |
| | 239863005 | Osteoarthritis of spinal facet joint |
| | 363215001 | Musculoskeletal system physical examination |
| | 279040009 | Mechanical low back pain |
| **Hypertension** | 59621000 | Essential hypertension |
| | 185720008 | Hypertension monitoring second letter |
| | 275944005 | Hypertension monitoring |
| | 185719002 | Hypertension monitoring first letter |
| | 401118009 | Hypertension annual review |
| | 185723005 | Hypertension monitoring telephone invite |
| | 1.06695E+15 | Hypertension monitoring SMS (short message service) text message first invitation |
| | 38341003 | Hypertensive disorder |
| | 185264001 | Seen in hypertension clinic |
| | 1.06696E+15 | Hypertension monitoring SMS (short message service) text message second invitation |
| | 38341003 | Hypertension |
| | 1.06694E+15 | Hypertension monitoring invitation SMS (short message service) text message |
| | 407567007 | Patient on maximal tolerated antihypertensive therapy |
| | 185721007 | Hypertension monitoring third letter |
| | 170577003 | Good hypertension control |
| | 161501007 | H/O: hypertension |
| | 310427004 | Hypertension monitoring invitation |
| | 38341003 | Hypertensive disease |
| | 401048005 | Hypertension six month review |
| | 417312002 | Suspected hypertension |
| **Urinary tract infection** | 307541003 | Lower urinary tract symptoms |
| | 314940005 | Suspected UTI (urinary tract infection) |
| | 4009004 | UTI - Lower urinary tract infection |

| | | |
|---|---|---|
| | 68566005 | UTI - Urinary tract infection |
| | 197927001 | Recurrent urinary tract infection |
| | 68566005 | Urinary tract infection |
| | 68566005 | Urinary tract infection, site not specified |
| | 721104000 | Sepsis due to urinary tract infection |
| | 197927001 | Recurrent urinary tract infections |
| | 307541003 | LUTS - Lower urinary tract symptoms |
| **Skin lesions** | 398838000 | Seborrhoeic keratosis |
| | 398838000 | Senile hyperkeratosis |
| | 95324001 | Skin lesion |
| | 201101007 | Actinic keratosis |
| | 201101007 | Solar keratosis |
| | 254666005 | Keratosis |
| | 315260001 | Subcutaneous component of pigmented skin lesion |
| | 315258003 | Crusting of pigmented skin lesion |
| **Diabetes** | 170777000 | Diabetic annual review |
| | 394675000 | O/E - Left diabetic foot at low risk |
| | 170742000 | Diabetic monitoring |
| | 394671009 | O/E - Right diabetic foot at low risk |
| | 185229005 | Seen in diabetic clinic |
| | 390850007 | O/E - no right diabetic retinopathy |
| | 170745003 | Diabetic on diet only |
| | 754121000000107 | Type II diabetic dietary review |
| | 44054006 | Type 2 diabetes mellitus |
| | 134395001 | Diabetic retinopathy screening |
| | 811981000000102 | Diabetes self-management plan agreed |
| | 313340009 | Seen in diabetic eye clinic |
| | 170746002 | Diabetic on oral treatment |
| | 887861000000105 | Diabetes Year of Care annual review |
| | 170744004 | Follow-up diabetic assessment |
| | 4855003 | Diabetic retinopathy |
| | 390834004 | Background diabetic retinopathy |
| | 170752001 | Has seen dietician - diabetes |
| | 390853009 | O/E - no left diabetic retinopathy |

| | | |
|---|---|---|
| | 415270003 | Referral to diabetes structured education programme |
| | 1.06691E+15 | Diabetes monitoring SMS (short message service) text message first invitation |
| | 700449008 | Non-diabetic hyperglycaemia |
| | 170763003 | Diabetic - good control |
| | 754081000000109 | Diabetic dietary review |
| | 394682001 | O/E - Right diabetic foot at moderate risk |
| | 700414001 | Education about diabetes and driving |
| | 185756006 | Diabetes monitoring first letter |
| | 284350006 | Diabetes mellitus diet education |
| | 705072004 | Diabetes monitoring invitation by short message service text messaging |
| | 408409007 | O/E - right eye background diabetic retinopathy |
| | 185757002 | Diabetes monitoring second letter |
| | 394681008 | O/E - Left diabetic foot at moderate risk |
| | 73211009 | Diabetes mellitus |
| | 703040004 | Agreeing on diabetes care plan |
| | 314194001 | Diabetic on insulin and oral treatment |
| | 755491000000100 | Diabetes structured education programme completed |
| | 170742000 | Diabetes management plan given |
| | 394725008 | Diabetes medication review |
| | 923461000000103 | Lifestyle education for diabetes |
| | 248161000000101 | Referral for diabetic retinopathy screening |
| | 185760009 | Diabetes monitoring telephone invite |
| | 407672007 | Seen in diabetic foot clinic |
| **Depression & Anxiety** | 198288003 | Anxiety state |
| | 191616006 | Recurrent depression |
| | 413973005 | Depression interim review |
| | 1.83956E+15 | PHQ9 total score 10-14 (moderate depression) |

| | | |
|---|---|---|
| | 197480006 | Anxiety disorder |
| | 231504006 | Mixed anxiety and depressive disorder |
| | 413974004 | Depression medication review |
| | 35489007 | Depression |
| | 197480006 | [X]Anxiety NOS |
| | 35489007 | Depressive disorder |
| | 70997004 | Mild anxiety |
| | 413972000 | Depression annual review |
| | 21897009 | Generalised anxiety disorder |
| | 445455005 | Generalized Anxiety Disorder 7 item score |
| | 48694002 | Anxiety |
| | 394924000 | Symptoms of depression |
| **Haemorrhoids & Constipation** | 70153002 | Haemorrhoids |
| | 14760008 | Constipation |
| | 14760008 | Constipation NOS |
| | 31499008 | Chronic constipation with overflow |
| | 70153002 | Piles - haemorrhoids |
| | 236069009 | Chronic constipation |
| **Asthma** | 394700004 | Asthma annual review |
| | 195967001 | Asthma |
| | 170635006 | Asthma not disturbing sleep |
| | 928451000000107 | Asthma monitoring invitation SMS (short message service) text message |
| | 527171000000103 | Patient has a written asthma personal action plan |
| | 170638008 | Asthma not limiting activities |
| | 406162001 | Asthma management |
| | 370208006 | Asthma never causes daytime symptoms |
| | 185731000 | Asthma monitoring call first letter |
| | 2.01003E+15 | Acute infective exacerbation of asthma |
| | 811921000000103 | Asthma self-management plan agreed |
| | 401182001 | Asthma monitoring by nurse |
| | 185732007 | Asthma monitoring call second letter |

| | | |
|---|---|---|
| | 394720003 | Asthma medication review |
| | 394701000 | Asthma follow-up |
| | 340911000000109 | Asthma trigger - pollen |
| | 394967008 | Suspected asthma |
| | 370202007 | Asthma causes daytime symptoms 1 to 2 times per month |
| | 810901000000102 | Asthma self-management plan review |
| | 161527007 | H/O: asthma |
| | 754061000000100 | Asthma review using Royal College of Physicians three questions |
| | 959401000000101 | Asthma monitoring SMS (short message service) text message first invitation |
| | 370218001 | Mild asthma |
| | 270442000 | Asthma monitoring check done |
| | 170636007 | Asthma never disturbs sleep |
| | 170658009 | Asthma never restricts exercise |
| | 401135008 | Health education - asthma |
| | 195967001 | Bronchial asthma |
| | 170656008 | Asthma sometimes restricts exercise |
| | 771901000000100 | Asthma causes night time symptoms 1 to 2 times per week |
| | 892301000000100 | Asthma management plan declined |
| | 185734008 | Asthma monitoring call third letter |
| | 31387002 | Exercise-induced asthma |
| | 370203002 | Asthma causes daytime symptoms 1 to 2 times per week |
| | 170634005 | Asthma disturbs sleep frequently |
| | 182729000 | Asthma control step 3 |
| | 401183006 | Asthma monitoring by doctor |
| | 370204008 | Asthma causes daytime symptoms most days |
| | 340901000000107 | Asthma trigger - exercise |
| | 170632009 | Asthma causing night waking |
| | 170657004 | Asthma severely restricts exercise |
| | 373899003 | Asthma daytime symptoms |
| | 866881000000101 | Chronic asthma with fixed airflow obstruction |

| | | |
|---|---|---|
| | 708038006 | Acute exacerbation of asthma |
| | 195967001 | Asthma NOS |
| | 185728001 | Attends asthma monitoring |
| | 233683003 | Pollen asthma |
| | 182726007 | Asthma control step 0 |
| | 182728008 | Asthma control step 2 |
| | 182727003 | Asthma control step 1 |
| | 370207001 | Asthma limits walking up hills or stairs |
| | 772051000000106 | Asthma limits activities most days |
| | 185730004 | Asthma monitor offer default |
| | 390877003 | Step up change in asthma management plan |
| | 370205009 | Asthma causes night symptoms 1 to 2 times per month |
| | 867171000000106 | No asthma trigger identified by subject |
| | 708373002 | Emergency asthma patient visit since last encounter |
| | 473391009 | Asthma never causes night symptoms |
| | 201051000000101 | Asthma trigger: animals |
| | 312453004 | Asthma - currently active |
| | 959421000000105 | Asthma monitoring SMS (short message service) text message second invitation |
| | 713701000000108 | Asthma monitoring administration |
| **Respiratory tract infection** | 50417007 | Lower respiratory tract infection |
| | 281794004 | Viral upper respiratory tract infection |
| | 275498002 | Respiratory tract infection |
| | 195742007 | Acute lower respiratory tract infection |
| **Rheumatoid arthritis** | 185227007 | Seen in rheumatology clinic |
| | 183526002 | Referred to rheumatologist |
| | 69896004 | Rheumatoid arthritis |
| | 847261000000104 | Rheumatoid arthritis annual review |
| | 239792003 | Seronegative rheumatoid arthritis |
| | 401192009 | Disease modifying antirheumatic drug monitoring |

| | | |
|---|---|---|
| | 364801000000104 | Seen in general practitioner disease modifying antirheumatic drug monitoring clinic |
| | 305749000 | Seen by rheumatology nurse specialist |
| | 306290009 | Referral to rheumatologist |

**Supplementary Table 6. Comparison between self-reported date of diagnosis with the first appearance of PD diagnosis in the EHR.** Patients without a coded diagnosis of PD were excluded from this analysis.To compare self-reported date of diagnosis in the baseline questionnaire with EHR record of PD diagnosis, observations (including records of symptoms, diagnoses, laboratory results and vital signs) and medication records of the first 200 participants were extracted. 7 participants were identified using their prescription records and had no coded diagnosis of PD in EHR. 1 participant was excluded due to their MSA-P diagnosis. The total number of participants included in the analysis was 192.

| **Lapse between self-reported date of diagnosis and first appearance of PD diagnosis in EHR** | **No. of patients (%)** |
|---|---|
| Within 1 year | 161 (83.9%) |
| Between 1-2 years | 14 (7.3%) |
| Between 2-3 years | 4 (2.1%) |
| Between 3-4 years | 5 (2.6%) |
| Between 4-5 years | 2 (1.0%) |
| More than 5 years | 6 (3.1%) |
| **Self-reported date earlier than first appearance of PD diagnosis in EHR** | 154 (80.2%) |
| **First appearance of PD diagnosis in EHR earlier than self-reported date of diagnosis** | 37 (19.3%) |
